# HLA-B Alleles with Shared Peptide Binding Specificities Define Global Risk of Cotrimoxazole-induced SCAR

**DOI:** 10.1101/2025.05.28.25328237

**Authors:** Yueran Li, Andrew Gibson, Hajirah N Saeed, Mohammad Ali Tahboub, Danmeng Li, David A Ostrov, Matthew S Krantz, Simon A Mallal, Eric Alves, Abha Chopra, Linda Choo, Roni P Dodiuk-Gad, Benjamin Kaffenberger, Aaron M Drucker, Michelle S Goh, Elizabeth Ergen, Robert Micheletti, Misha Rosenbach, Michelle D Martin-Pozo, Rama Gangula, Elizabeth A Williams, Alexis Yu, April O’Connor, Kelby Mahan, James T Kwan, Derek Metcalfe, Ramy Rashad, Swapna S Shanbhag, Sarah Pedretti, Phuti Choshi, Tafadzwa Chimbetete, Rose Selim, Ian James, Jason A Trubiano, Rannakoe Lehloenya, Jonny G Peter, Elizabeth J Phillips, the SJS Survivor Study

## Abstract

**Background:** Co-trimoxazole is a leading global cause of severe cutaneous adverse drug reactions (SCAR) including Stevens-Johnson syndrome/toxic epidermal necrolysis (SJS/TEN) and drug reaction with eosinophilia and systemic symptoms (DRESS). Co-trimoxazole-induced SCAR are associated with HLA class I alleles including HLA-B*13:01 and HLA-B*38:02 in Southeast Asian (SEA) populations. However, the global generalizability of these associations is unknown but critical for population-appropriate risk stratification and diagnosis.

**Objective:** To determine HLA risk factors associated with co-trimoxazole-induced SJS/TEN and DRESS in populations from the United States (US) and South Africa (SA).

**Methods:** We performed high-resolution HLA typing on dermatologist-adjudicated co-trimoxazole-induced SCAR patients in the US (n=63) and SA (n=26) compared to population controls. Peptide binding and docking analyses were performed using MHCcluster2.0 and CB-Dock2.

**Results:** In a multiple logistic regression model, HLA-B*44:03 (Pc<0.001, OR: 4.08), HLA-B*38:01 (Pc<0.001, OR: 5.66), and HLA-C*04:01 (Pc=0.003, OR: 2.50) were independently associated with co-trimoxazole-induced SJS/TEN in the US. HLA-B*44:03 was also associated with co-trimoxazole-induced DRESS in SA (Pc=0.019, OR: 10.69). Distinct HLA-B variants with shared peptide binding specificities (SPBS) and HLA-C*04:01 identified 94% and 78% of co-trimoxazole-induced SJS/TEN and DRESS in the US, respectively. The SEA risk allele HLA-B*13:01, with SPBS to HLA-B*44:03, was identified in just 1/63 US SCAR patients.

**Conclusion:** HLA alleles with SPBS to SEA-related risk alleles including HLA-B*44:03 (SPBS with HLA-B*13:01) and HLA-B*38:01 (SPBS with HLA-B*38:02) but also HLA-C*04:01 predisposed to co-trimoxazole-induced SCAR in the US and SA. These findings provide biological plausibility and strategies for global risk prediction and diagnosis of co-trimoxazole-induced SCAR.

**HIGHLIGHTS BOX:** *What is already known about this topic?:* HLA alleles including HLA-B*13:01 and HLA-B*38:02 are risk factors for co-trimoxazole-induced SCAR in Asian populations. However, the generalizability of these associations to other global populations is unknown but critical for population-appropriate risk stratification and diagnosis.

*What does this article add to our knowledge?:* HLA alleles with shared peptide binding specificities (SPBS) to Asian-related risk alleles including HLA-B*44:03 (SPBS with HLA-B*13:01) and HLA-B*38:01 (SPBS with HLA-B*38:02) but also HLA-C*04:01 predisposed to co-trimoxazole-induced SCAR in the US and South Africa.

*How does this study impact current management guidelines?:* HLA alleles previously associated with co-trimoxazole-induced SCAR do not identify risk across populations. However, HLA alleles with SPBS provide biological plausibility and strategies for global and population-appropriate clinical risk stratification and diagnosis of cotrimoxazole-induced SCAR.

## Introduction

Co-trimoxazole is an antibiotic used globally for community- and hospital-acquired bacterial infections and opportunistic infection prophylaxis in persons living with HIV (PLWH) [1]. However, co-trimoxazole is also a common cause of T-cell-mediated severe cutaneous adverse reactions (SCAR), including drug reaction with eosinophilia and systemic symptoms (DRESS) and Stevens-Johnson Syndrome/toxic epidermal necrolysis (SJS/TEN) with mortalities of 3-10% and 15-70%, respectively [2–4]. Importantly, the rarity of SJS/TEN (1-5 per million patients/annum) and DRESS (1/1,000-10,000 per drug course) makes it challenging to study epidemiological and genetic risk [5].

A recent multi-country case-control study including 151 co-trimoxazole-induced SCAR patients and 4631 population controls from Taiwan, Thailand, and Malaysia identified a strong and reproducible association between co-trimoxazole-induced DRESS and HLA-B*13:01 carried by 85% of patients [6]. In 2024, a subsequent meta-analysis of six predominantly Southeast Asian (SEA) studies of co-trimoxazole-induced SCAR identified multiple HLA associations including HLA-A*11:01, HLA-B*13:01, HLA-B*15:02, HLA-B*38:02, and HLA-C*08:01 [7]. Despite differential associations for SJS/TEN (HLA-B*38:02, HLA-B*15:02) and DRESS (HLA-A*68:01, HLA-B*39:01), HLA-B*13:01 was associated with both SJS/TEN and DRESS [7]. However, while HLA-B*13:01 is carried in 2-10% of the SEA population [8], it is rare amongst those with European (0-1.2%) and African ancestry (<0.04%) [9] and its relevance in other global populations is unknown but will be critical for population-appropriate clinical risk stratification and diagnosis.

The United States (US) represents a large geographical population with high exposure to co-trimoxazole. Further, in South Africa (SA), with 70% of the global HIV burden [10], co-trimoxazole is recommended prophylactically in PLWH [1]. Thus, our objective was to investigate HLA associations with co-trimoxazole-induced SCAR using prospectively-collected and geographically-matched co-trimoxazole-induced SCAR survivors and population-based controls in the US and SA.

## Methods

### Co-trimoxazole-induced DRESS and SJS/TEN cases

US SCAR patients were recruited through 1) Prospective identification through outpatient and inpatient facilities at Vanderbilt University Medical Center (VUMC), Nashville. 2) A prospective cohort of SJS/TEN patients from Massachusetts Eye and Ear Institute (MEEI), Boston. 3) The SJS Survivor Study: a community-based study recruiting SJS/TEN survivors for whom medical records were redacted to determine eligibility and a DNA sample obtained. From SA, SCAR patients were recruited prospectively at Groote Schuur Hospital, University of Cape Town, and Nelson Mandela Academic Hospital, Walter Sisulu University. An additional Australian population was identified from the Australasian Registry for (AUS)-SCAR [11]. In the US and SA, 76% and 50% of SCAR patients were female, respectively, which may reflect a higher incidence of SCAR in women than men, however, we could not exclude the potential for more women to participate in survivor studies. SCAR phenotypes were adjudicated by independent dermatologists with probable or definite co-trimoxazole-induced SCAR with ALDEN score ≥4 for SJS/TEN [12], and a Naranjo score ≥5 and RegiSCAR ≥4 for DRESS [13, 14].

### Ethical statement

All samples and data were obtained with informed consent under institutional review board (IRB) approvals from VUMC (IRB 131836, 150754, 210027, 191350, 171900), MEEI (IRB 654182), Walter Sisulu University (HREC 056/2020), University of Cape Town (HREC R031/2018, 500/2018), Austin Health (HREC 50791/Austin-2019), and Murdoch University (HREC 2011/056, 2017/246, 2019/153). Samples and data were de-identified.

### Large geographic population control cohorts in the US and SA

Our US control population was BioVU; a de-identified health record-linked DNA database [15] including genomic data of 94,489 individuals identified by genetic race as White (79%), Black (17%), Asian (3%), and other (1%). For geographical concordance, this was consistent with US SCAR patients (White, 78%; Black, 13%; Asian, 6%) providing a large population ethnicity-matched control. For SA, we utilized two published databases. First, 159 individuals from the Western Cape [16]. Second, 192 healthy individuals from Cape Town [17, 18]. Aligned with the predominant self-identified ethnicities of SA SCAR patients, the Western Cape population is predominantly of Black African and Cape Mixed or South African “Coloured” ancestry [17].

### HLA genotyping

For US, SA, and Australian cases high-resolution HLA class I (HLA-A,B,C) and II (HLA-DQA1,-DQB1,-DRB1,-DPB1) typing was performed at the National Association of Testing Authorities (NATA)- and American Society for Histocompatibility and Immunogenetics (ASHI)-accredited genomics laboratories at Murdoch University and the Immunogenomics and Microbial Genetics Single-Cell Technologies core at VUMC on DNA extracted from whole blood EDTA, cryopreserved PMBC, or saliva collected by Oragene-500 kits. HLA sequencing was performed using the V3 600-cycle kit (2X300bp reads) on the Illumina MiSeq as previously described [19].

### HLA association and peptide binding analyses

Carriage of HLA alleles in US or SA SCAR patients were compared to the US BioVU or SA population controls, respectively, using R (v4.3.2) and SPSS for Fisher’s exact test and logistic regression with Bonferroni correction. Joint associations of alleles were assessed for individual significance using multiple case-control logistic regression. To assess similar peptide binding of HLA class I alleles without knowledge of immunogenic peptide(s); we utilized MHCcluster 2.0 [20]. HLA supertypes are based on overlapping peptide binding and we aligned alleles to the B44 supertype using published reference studies [21–23].

### Molecular docking

HLA structures were selected from the Protein Data Bank for molecular docking studies of HLA-B*13:01 (ID:7YG3), HLA-B*44:03 (ID:4JQX), and HLA-C*04:01 (ID:1QQD). Structural models of HLA-B*38:01 were generated using SWISS-MODEL [24]. Docking simulations were conducted with AutoDock Vina [25] and CB-Dock2 [26] to estimate affinity and binding positions within the HLA antigen-binding cleft. The 3D structures of drug antigens including sulfamethoxazole (SMX), trimethoprim (TMP), sulphanilamide, and dapsone, were obtained from Drugbank (https://go.drugbank.com/). Drug antigens are shown as sticks (white, carbon; blue, nitrogen; red, oxygen; yellow, sulfur). Docking scores were reported and represent the predicted energy of binding Δ values in kcal/mol. To model drug antigen docking in the presence of peptides, we used known HLA-related virotopes and TCRModel.

## Results

### Co-trimoxazole-SCAR patients in the US and SA

In the US we identified 54 patients with co-trimoxazole-induced SJS/TEN and 9 patients with co-trimoxazole-induced DRESS. In SA, we identified 15 patients with co-trimoxazole-induced SJS/TEN and 11 patients with co-trimoxazole-induced DRESS, of which 73% and 91%, respectively, were PLWH. Patients presented with a range of latency, severity, and mortality (Table E1).

### HLA-B*44:03, HLA-B*38:01, and HLA-C*04:01 are risk factors for co-trimoxazole-SJS/TEN in the US

The US population was powered towards identifying risk for SJS/TEN. Patient HLA typing and univariate analyses are listed in Tables E2-3. Most significantly, a new association was found between HLA-B*44:03 and co-trimoxazole-induced SCAR (Table 1; odds ratio (OR): 3.75; 95% confidence interval (CI): 2.04-6.61; Pc=0.002) and SJS/TEN (OR: 4.68; CI: 2.50-8.47; Pc<0.001). A significant association was also identified between HLA-B*38:01 and co-trimoxazole-induced SCAR (Table 1; OR: 4.47; CI: 1.57-10.36; P=0.003) and SJS/TEN (OR: 4.33; CI: 1.35-10.84; P=0.008) although only prior to correction. However, HLA-B*38 is previously reported in a small European study [27]. Finally, we found a new association between HLA-C*04:01 and co-trimoxazole-induced SJS/TEN (Table 1; OR: 2.95; CI: 1.65-5.22; Pc=0.021). HLA-B*38:01, HLA-B*44:03, and HLA-C*04:01 were carried by 10%, 29%, and 40% of SCAR patients, respectively, but just 2%, 10%, and 21% of BioVU controls. Alleles were in Hardy Weinberg equilibrium (P>0.05) (Table E4).

**Table 1.** HLA alleles associated with Co-trimoxazole-induced SCAR in the US. HLA alleles associated with SCAR, SJS/TEN, and DRESS in the US patient versus BioVU population cohorts. P values were calculated using Fisher’s exact test with Bonferroni correction. Significant outcomes (Pc<0.05) are highlighted red. Subsets with <3 patients were not analyzed. *OR, odds ratio; C.I, confidence internal*.

| HLA allele | SCAR Phenotype | Case N(%) | Population Control N(%) | OR (95% C.I.) | Fisher's P value | Bonferroni $P_c$ |
| --- | --- | --- | --- | --- | --- | --- |
| HLA-B*38:01 | Co-TSCAR (n=63) | 6 (10%) | 2173 (2%) | 4.47 (1.57, 10.36) | **0.003** | 0.341 |
|  | Co-TSJS/TEN (n=54) | 5 (9%) |  | 4.33 (1.35, 10.84) | **0.008** | 0.652 |
|  | Co-TDRESS (n=9) | 1 (11%) |  | -- | -- | -- |
| HLA-B*44:03 | Co-TSCAR (n=63) | 18 (29%) | 9114 (10%) | 3.75 (2.04, 6.61) | **<0.001** | **0.002** |
|  | Co-TSJS/TEN (n=54) | 18 (33%) |  | 4.68 (2.50, 8.47) | **<0.001** | **<0.001** |
|  | Co-TDRESS (n=9) | 0 |  | -- | -- | -- |
| HLA-C*04:01 | Co-TSCAR (n=63) | 25 (40%) | 20153 (21%) | 2.43 (1.40, 4.13) | **<0.001** | 0.117 |
|  | Co-TSJS/TEN (n=54) | 24 (44%) |  | 2.95 (1.65, 5.22) | **<0.001** | **0.021** |
|  | Co-TDRESS (n=9) | 1 (11%) |  | -- | -- | -- |

While 7 US SCAR patients carried both HLA-B*44:03 and HLA-C*04:01 these were not predicted to be haplotypic (SCAR patients, R^2^=0.038; BioVU, R^2^=0.033; Figure E1), indicating independent risk factors. To confirm, we performed case-control logistic regression using covariates identified for HLA risk (Table 2), including HLA-B*38:01, which is closely related to HLA-B*38:02 (differing by only three amino acids at positions 116, 156 and 163); a risk factor in SEA populations. Here, HLA-B*44:03, HLA-B*38:01, and HLA-C*04:01 were shown as independent risk factors for co-trimoxazole-induced SCAR (HLA-B*44:03: OR: 3.38, CI: 1.88-5.83, Pc<0.001; HLA-B*38:01: OR: 5.55, CI: 2.13-11.97, Pc<0.001; HLA-C*04:01: OR: 2.13, CI: 1.25-3.55, Pc=0.012) and SJS/TEN (HLA-B*44:03: OR: 4.08, CI: 2.23-7.21, Pc<0.001; HLA-B*38:01: OR: 5.66, CI: 1.95-13.04, Pc<0.001; HLA-C*04:01: OR: 2.50, CI: 1.43-4.33, Pc=0.003) (Table 2).

**Table 2.**
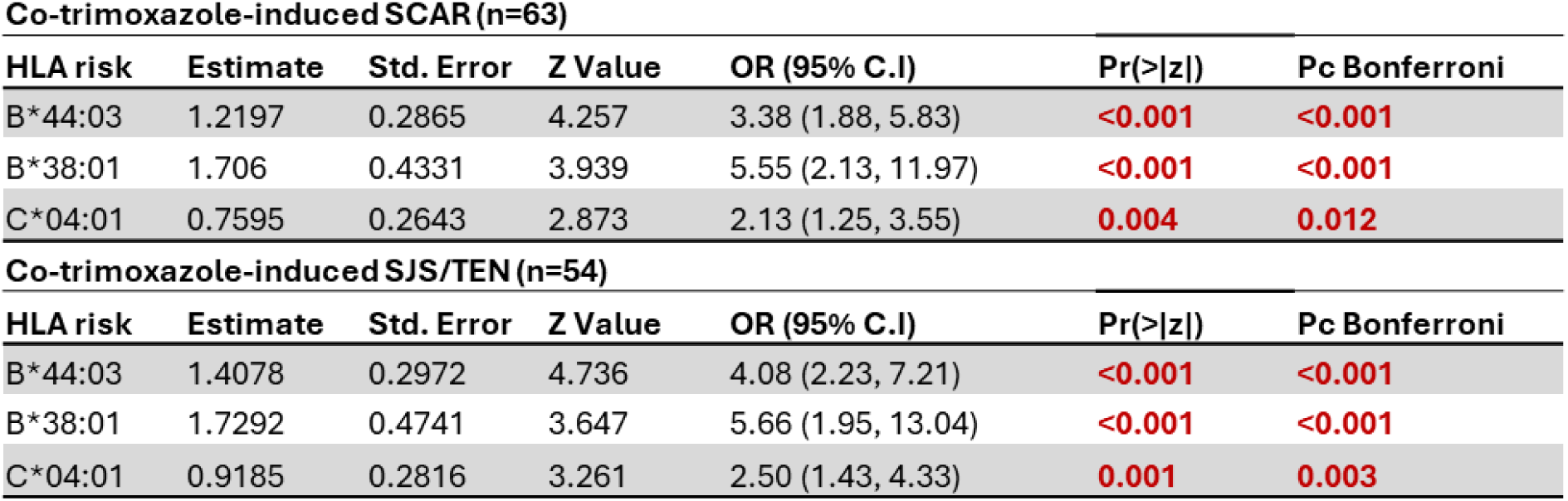
Conditional logistic regression analyses for co-trimoxazole-induced SCAR and SJS/TEN in the US population. HLA-B*38:01, HLA-B*44:03, and HLA-C*04:01 were included as co-variates with Bonferroni correction. Significant outcomes (Pc<0.05) are highlighted red. *OR, odds ratio; C.I, confidence interval; Pr, probability*.

| **Co-trimoxazole-induced SCAR (n=63)** |  |  |  |  |  |  |
| --- | --- | --- | --- | --- | --- | --- |
| **HLA risk** | **Estimate** | **Std. Error** | **Z Value** | **OR (95% C.I.)** | **Pr(>\|z\|)** | **Pc Bonferroni** |
| B*44:03 | 1.2197 | 0.2865 | 4.257 | 3.38 (1.88, 5.83) | **<0.001** | **<0.001** |
| B*38:01 | 1.706 | 0.4331 | 3.939 | 5.55 (2.13, 11.97) | **<0.001** | **<0.001** |
| C*04:01 | 0.7595 | 0.2643 | 2.873 | 2.13 (1.25, 3.55) | **0.004** | **0.012** |
| **Co-trimoxazole-induced SJS/TEN (n=54)** |  |  |  |  |  |  |
| **HLA risk** | **Estimate** | **Std. Error** | **Z Value** | **OR (95% C.I.)** | **Pr(>\|z\|)** | **Pc Bonferroni** |
| B*44:03 | 1.4078 | 0.2972 | 4.736 | 4.08 (2.23, 7.21) | **<0.001** | **<0.001** |
| B*38:01 | 1.7292 | 0.4741 | 3.647 | 5.66 (1.95, 13.04) | **<0.001** | **<0.001** |
| C*04:01 | 0.9185 | 0.2816 | 3.261 | 2.50 (1.43, 4.33) | **0.001** | **0.003** |

### Shared binding groups predispose co-trimoxazole-induced SJS/TEN in the US

HLA-B*13:01 and HLA-B*40:01 are associated with co-trimoxazole-induced SCAR in SEA populations [6, 28] and have shared peptide binding specificity (SPBS). To define whether SPBS defines risk across populations, we grouped HLA-B alleles of US SCAR patients by SPBS (Figure 1A). Four HLA-B clusters included alleles carried by ≥10 SCAR patients. The most prevalent allele was used as a reference, resulting in ‘B*44:03-like’, ‘B*38:01-like’, ‘B*35:02-like’, and ‘B*35:01-like’ clusters (Figure 1A). Importantly, alleles in both the ‘B*44:03-like’ and ‘B*38:01-like’ clusters also overlapped with the B44 supertype (Figure 1B, Table E5) defined by Sidney [22, 23] and Doytchinova [21].

**Figure 1.**
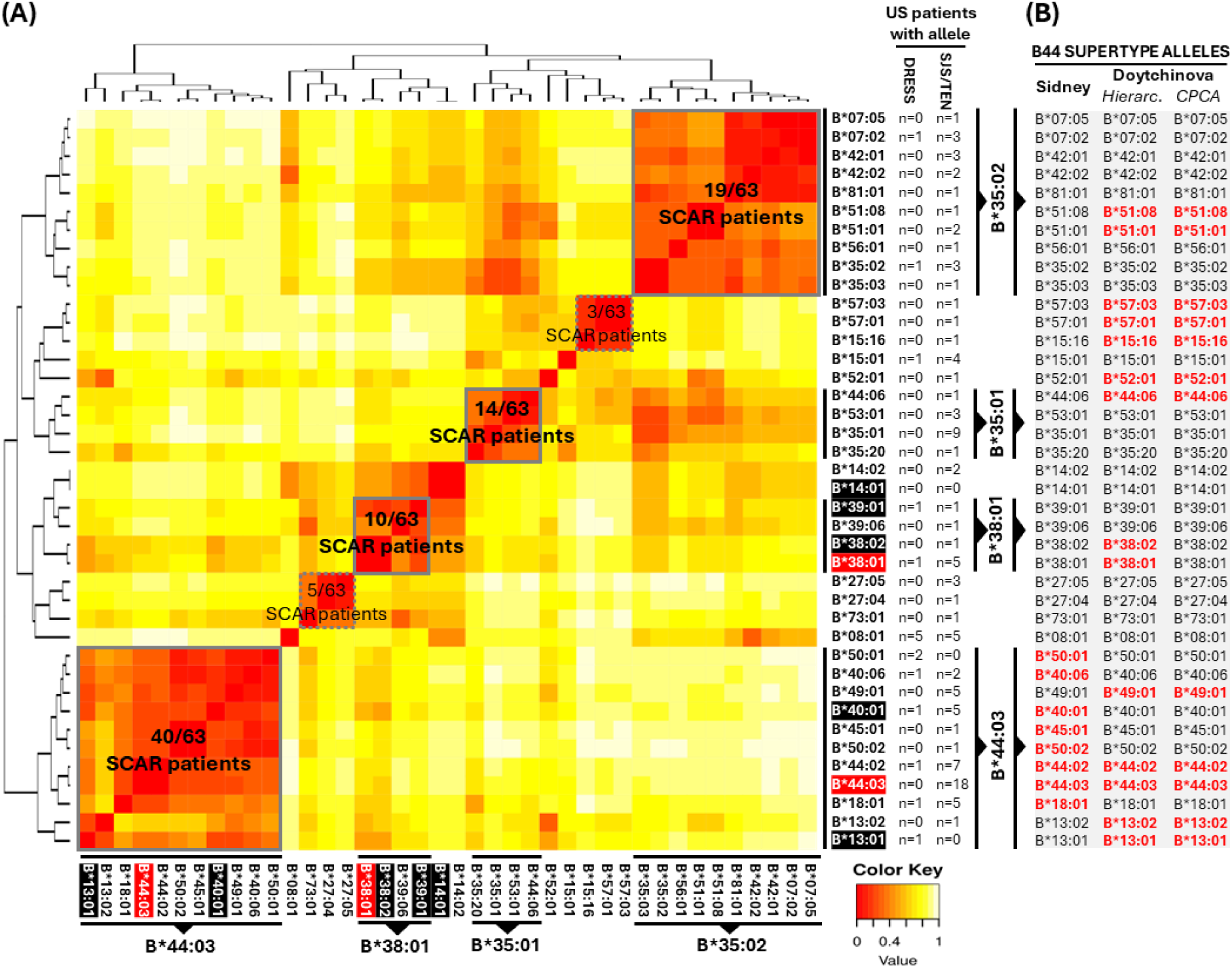
HLA-B alleles in US co-trimoxazole-SCAR patients share peptide binding and supertype specificities. HLA-B alleles cluster by shared **(A)** peptide binding (high to low, red to yellow) and **(B)** B44 supertype specificities. Previously defined associations are highlighted black and those reported here in red. HLA-B*14:01 is not expressed in the US cohort but is reported in literature. *CPCA, consensus principal component analyses*.

First, we investigated an association between co-trimoxazole-SCAR in the US and HLA-B alleles with SPBS. The ‘B*44:03-like’ cluster encompassed HLA-B*44:03, HLA-B*13:01, HLA-B*40:01, HLA-B*45:01, HLA-B*50:01, HLA-B*50:02, HLA-B*44:02, HLA-B*40:06, HLA-B*49:01, and HLA-B*18:01 (Figure 1A). While HLA-B*44:03 was carried by 29% of SCAR patients and 10% of controls, the ‘B*44:03-like’ cluster identified 63% of SCAR patients and 49% of controls. Only 1/63 (1.6%) SCAR patients carried the SEA-related allele HLA-B*13:01 (Table E2). The ‘B*44:03-like’ cluster was significantly associated with co-trimoxazole-induced SCAR (Pc=0.005, OR: 2.23, CI: 1.33-3.84) and SJS/TEN (Pc<0.001, OR: 2.70, CI: 1.56-4.86) (Table 3A). Only 1/9 DRESS patients carried HLA-B*44:03, however 5/9 patients carried ‘B*44:03-like’ cluster alleles and this was significantly associated with DRESS (Table 3A; Pc=<0.001, OR: 12.84, CI: 4.70-49.02).

**Table 3.** Shared peptide binding and supertype specificities are associated with broad population risk for co-trimoxazole-induced SCAR in the US. HLA-B alleles with shared **(A)** peptide binding or **(B)** B44 supertype specificities in the US SCAR patient and population cohorts. P values calculated using Fisher’s exact test with Bonferroni correction. Significant outcomes (Pc<0.05) are highlighted red. Subsets with <3 patients were not analyzed. *OR, odds ratio; C.I, confidence interval*.

| **(A) HLA-B Clustered alleles based on peptide binding specificity** | **Reaction phenotype** | **Cases N(%)** | **Population control (N=94489) N(%)** | **OR (95% C.I.)** | **Bonferroni $P_c$** |
| --- | --- | --- | --- | --- | --- |
| **B\*44:03-like cluster** | Co-T SCAR (n=63) | 40 (63%) | 46144 (49%) | 2.23 (1.33, 3.84) | **0.005** |
|  | Co-T SJS/TEN (n=54) | 35 (65%) |  | 2.70 (1.56, 4.86) | **<0.001** |
|  | Co-T DRESS (n=9) | 5 (56%) |  | 12.84 (4.70, 49.02) | **<0.001** |
| **B\*38:01-like cluster** | Co-T SCAR (n=63) | 10 (16%) | 5864 (6%) | 1.71 (0.60, 3.98) | 0.551 |
|  | Co-T SJS/TEN (n=54) | 8 (15%) |  | 1.97 (0.69, 4.63) | 0.447 |
|  | Co-T DRESS (n=9) | 2 (22%) |  | -- | -- |
| **B\*35:02-like cluster** | Co-T SCAR (n=63) | 19 (30%) | 35673 (38%) | 1.42 (0.90, 2.25) | 0.378 |
|  | Co-T SJS/TEN (n=54) | 17 (31%) |  | 1.69 (1.05, 2.74) | 0.091 |
|  | Co-T DRESS (n=9) | 2 (22%) |  | -- | -- |
| **B\*35:01-like cluster** | Co-T SCAR (n=63) | 14 (22%) | 13926 (15%) | 1.77 (0.92, 3.21) | 0.267 |
|  | Co-T SJS/TEN (n=54) | 14 (26%) |  | 2.17 (1.11, 4.02) | 0.052 |
|  | Co-T DRESS (n=9) | 0 |  | -- | -- |
| **(B) HLA-B44 supertypes** | **Reaction phenotype** | **Cases N(%)** | **Population control (N=94489) N(%)** | **OR (95% C.I.)** | **Bonferroni $P_c$** |
| **B44 supertype (Sidney)** | Co-T SCAR (n=63) | 37 (59%) | 25489 (26%) | 3.85 (2.27, 6.63) | **<0.001** |
|  | Co-T SJS/TEN (n=54) | 31 (57%) |  | 3.65 (2.06, 6.55) | **<0.001** |
|  | Co-T DRESS (n=9) | 6 (67%) |  | 5.41 (1.16, 33.49) | **0.044** |
| **B44 Hierachial (Doytchinova)** | Co-T SCAR (n=63) | 40 (64%) | 47239 (48%) | 1.74 (1.02, 3.04) | 0.123 |
|  | Co-T SJS/TEN (n=54) | 38 (70%) |  | 2.38 (1.29, 4.56) | **0.012** |
|  | Co-T DRESS (n=9) | 2 (22%) |  | -- | -- |
| **B44 CPCA (Doytchinova)** | Co-T SCAR (n=63) | 34 (54%) | 44916 (50%) | 1.29 (0.77, 2.20) | 0.679 |
|  | Co-T SJS/TEN (n=54) | 33 (61%) |  | 1.73 (0.97, 3.15) | 0.158 |
|  | Co-T DRESS (n=9) | 1 (11%) |  | -- | -- |

The ‘B*38:01-like’ cluster included HLA-B*38:01 and HLA-B*38:02, representing reported risk alleles [27, 29], and HLA-B*39:01, also identified as a weak association in SEA populations [6]. However, the ‘B*38:01-like’ cluster was not significantly associated with co-trimoxazole-induced SCAR in our study (Table 3A). We were likely underpowered to show this given independent associations with 3/4 alleles in this cluster. Remaining clusters were not associated with co-trimoxazole-SCAR.

Second, we investigated associations with the B44 supertype. The hierarchical clustering-defined B44 supertype including HLA-B*38:01 and HLA-B*38:02 was associated with co-trimoxazole-induced SJS/TEN (Pc=0.011, OR: 2.38, CI: 1.29-4.56), and the Sidney B44 supertype, which only includes alleles in our cohort from the ‘B*44:03-like’ cluster, was significantly associated with co-trimoxazole-induced SCAR (Pc=<0.001, OR: 3.85, CI: 2.27-6.63), SJS/TEN (Pc<0.001, OR: 3.65, CI: 2.06-6.55), and DRESS (Pc=0.044, OR: 5.41, CI: 1.16-33.49) (Table 3B). Overall, the ‘B*44:03-like’ cluster, ‘B*38:01-like’ cluster, and HLA-C*04:01 identified 51/54 (94%) SJS/TEN and 7/9 (78%) DRESS patients (Table E2).

We also identified patients with SJS/TEN (n=7) and DRESS (n=4) caused by sulfasalazine, which is an anti-inflammatory drug cleaved in the gut to the sulfanilamide derivative antibiotic sulfapyridine. Importantly, 10/11 patients carried HLA-B*38:01, HLA-B*44:03, or HLA-C*04:01 (Table E2); the same risk alleles identified for co-trimoxazole suggesting shared HLA restriction across sulfonamide antibiotics.

### Molecular docking of drug antigens on HLA-B*38:01, HLA-B*44:03, and HLA-C*04:01

Co-trimoxazole is comprised of SMX and TMP. Interestingly, when the association with HLA-B*13:01 was first described, without a defined structure for HLA-B*13:01, the known structure of HLA-B*44:03 was used to model docking with SMX due to high (>96%) sequence homology [6]. The protein structure of HLA-B*13:01 is since validated [30] and we model HLA-B*44:03, HLA-B*13:01, and HLA-C*04:01 with TMP, SMX, and the immunogenic metabolite nitroso-sulfamethoxazole (SMX-NO) (Figure 2A-B) [31–33]. We also model sulphanilamide and dapsone, and nevirapine, due to similar reported associations with HLA-B*13:01 [34, 35] and HLA-C*04:01 [36, 37], respectively. Predominantly, for HLA-B*13:01 and HLA-B*38:01, compounds bound to the A/B pocket (residues 7, 63, 66, 167), while for HLA-B*44:03 and HLA-C*04:01, compounds bound in pockets C/D (residues 9, 70, 97, 99, 156). These results demonstrate consistent docking of SMX, TMP, and SMX-NO within the antigen-binding clefts of the same HLA molecule; however, multiple docking orientations and poses are observed across alleles (Figure 2A-B). The highest predicted docking to HLA-C*04:01 was with nevirapine, SMX, and SMX-NO (Figure 2C). Across HLA-B*44:03 and HLA-B*13:01, the highest predicted binding was for SMX and SMX-NO, with a weaker score for TMP (Figure 2C). These data indicate that, like HLA-B*13:01, HLA-C*04:01 and HLA-B*44:03 are associated with reactions driven by the sulfonamide antibiotic moiety. Interestingly, HLA-B*44:03 binding scores were weaker for dapsone. Thus, unlike HLA-B*13:01, which is associated with co-trimoxazole- and dapsone-induced SCAR in SEA populations [6, 35], HLA-B*44:03 may be more structurally selective, warranting future studies as some reports have suggested limited clinical cross-reactivity between dapsone and co-trimoxazole [38].

**Figure 2.**
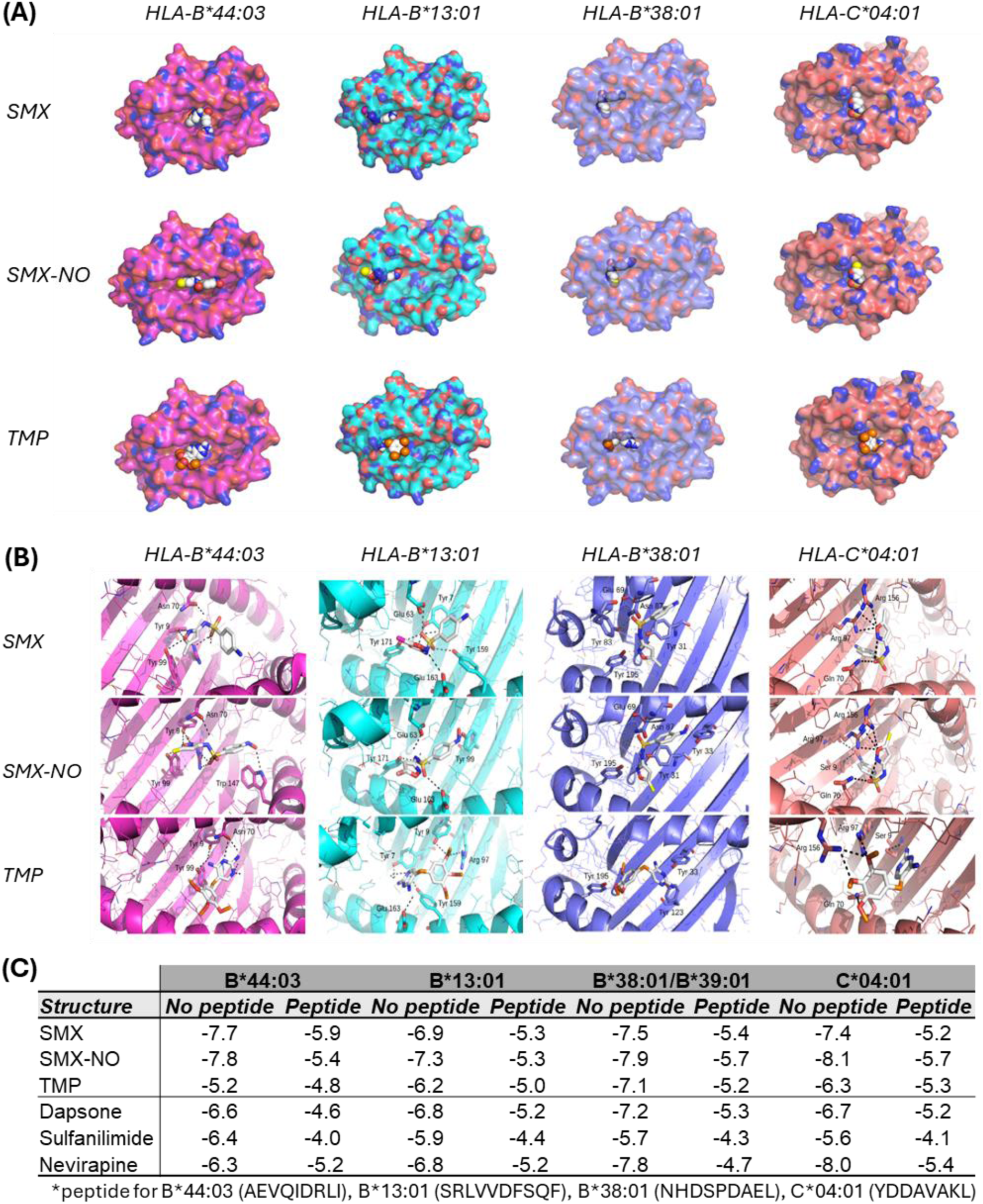
HLA-B*44:03, HLA-B*13:01, HLA-B*38:01, and HLA-C*04:01 exhibit enhanced binding affinity for sulfamethoxazole compared to trimethoprim. **(A)** Drug antigen docking in the presence of HLA-related peptides. **(B)** Zoomed views of drug antigens docked to HLA-B*44:03, HLA-B*13:01, HLA-B*38:01, and HLA-C*04:01. **(C)** Docking scores represent the predicted binding energy (kcal/mol). *SMX, Sulfamethoxazole; TMP, Trimethoprim; SMX-NO, Sulfamethoxazole-nitroso*.

### Predictive value of HLA-B*44:03, HLA-B*38:01, and HLA-C*04:01 in the US

We calculated the positive predictive value (PPV), negative predictive value (NPV), and the number needed to be HLA tested (NNT) to prevent one case in the US (Table E6). While SCAR is reported in 0.9% of patients administered co-trimoxazole in Taiwan [6], we use a conservative 0.1%; aligned with the incidence of DRESS [4]. HLA-B*38:01 provided the highest PPV (0.4%) but the lowest sensitivity (9.5%), with an NNT of 9,932. In contrast, the PPV of HLA-B*44:03 and HLA-C*04:01 was low (0.2-0.3%) but each had higher sensitivity of 28.6% and 39.7%, respectively, providing a theoretical NNT of 3,311 and 2,384. Using BioVU controls, screening for HLA-B*38:01, HLA-B*44:03, and HLA-C*04:01 would identify 2%, 10%, and 21% of the US population, respectively. The NNT was improved by combining risk alleles, with screening for HLA-B*38:01, HLA-B*44:03, and HLA-C*04:01 reducing the NNT to 1,453 and identifying 31% of BioVU controls. Further, the combination of the ‘B*44:03-like’ cluster, ‘B*38:01-like’ cluster, and HLA-C*04:01 reduced the NNT to 1,010 (Table E6). Using BioVU, this would exclude 63% of the US population and would not represent a feasible screening approach until additional risk factors are discovered to improve predictivity. However, this approach could be implemented to help with clinical risk stratification, diagnosis, and retrospective drug causality assessment.

### HLA-B*44:03 is associated with co-trimoxazole-induced SCAR in other populations

We investigated risk in SA by comparing the HLA carriage of population controls (n=351) to patients with co-trimoxazole-induced DRESS (n=11) and SJS/TEN (n=15) (Tables E7-8). HLA-B*44:03 was the only allele significantly associated with co-trimoxazole-induced DRESS (Pc=0.019, OR: 10.69, CI: 2.57-46.75) and carried by 55% of SA DRESS patients but 10% of controls (Table 4). ‘B*44:03-like’ cluster alleles were also identified in SA SCAR patients, including HLA-B*45:01 and HLA-B*18:01 (Figure 3, Table E7). Further, although HLA-B*38:01 was not carried by SA co-trimoxazole-induced SCAR patients, HLA-B*15:10 has SPBS and was carried by 3 SCAR patients (Figure 3). In contrast, no SA patients carried the SEA-related risk allele HLA-B*13:01. The association with HLA-B*44:03 highlights a shared predisposition to both co-trimoxazole-induced SJS/TEN (US) and DRESS (SA). Further, in support of global risk, and although currently underpowered for independent study, we also report ‘B*44:03-like’ cluster alleles carried by 5/6 SJS/TEN and 4/6 DRESS patients in Australia (Table E9, Figure E2).

**Figure 3.**
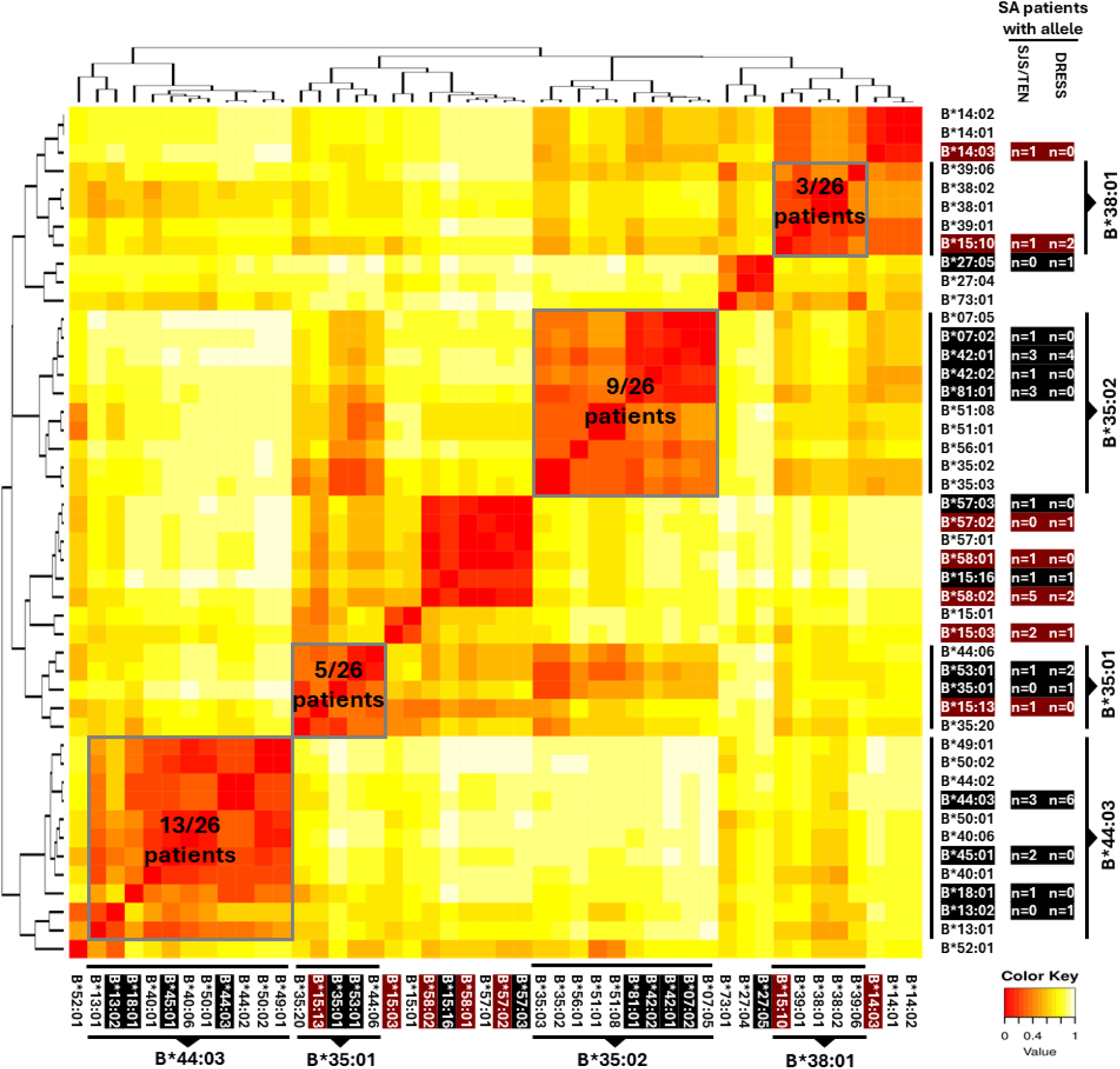
Integration of HLA-B alleles with shared peptide binding specificities in co-trimoxazole-SCAR patients in South Africa with the US. HLA-B alleles clustered by shared peptide binding (high to low, red to yellow). Alleles present in both US and South African cohorts are highlighted black, and those unique to the South African cohort highlighted red. HLA-B*08:01 was not linked to any other allele and removed to aid prediction.

**Table 4.** HLA alleles associated with Co-trimoxazole-induced SCAR in South Africa. HLA alleles associated with SCAR, SJS/TEN, and DRESS in the South African patient versus control cohorts. P values were calculated using Fisher’s exact test with Bonferroni correction. Significant outcomes (Pc<0.05) are highlighted red. Subsets with <3 patients were not analyzed. *OR, odds ratio; C.I, confidence internal*.

| HLA allele | Reaction phenotype | Case N(%) | Population control N(%) | OR (95% C.I) | Fisher's P value | Bonferroni $P_c$ |
| --- | --- | --- | --- | --- | --- | --- |
| B*44:03 | SCAR (N=26) | 9 (35%) | 35 (10%) | 4.75 (1.73, 12.31) | **0.001** | 0.066 |
|  | DRESS (N=11) | 6 (55%) |  | 10.69 (2.57, 46.75) | **<0.001** | **0.019** |
|  | SJS/TEN (N=15) | 3 (20%) |  | 2.25 (0.39, 8.91) | 0.196 | -- |

## Discussion

The SEA-related co-trimoxazole-induced SCAR risk allele HLA-B*13:01 is not commonly found in US or SA SCAR patients. Instead, we identified alleles with SPBS including HLA-B*44:03 (SPBS with SEA risk allele HLA-B*13:01 [6, 7, 28]) and HLA-B*38:01 (SPBS with SEA risk allele HLA-B*38:02 [7, 28]) but also HLA-C*04:01 that predisposed to co-trimoxazole-induced-SCAR and identified 92% and 73% of patients in the US and SA, respectively.

Our findings underscore the importance of population-specific studies, as diverse HLA alleles previously associated with co-trimoxazole-induced SCAR in SEA populations [7] were infrequent amongst patients in the US. Specifically, no US patients carried HLA-B*15:02, two carried HLA-B*39:01, and HLA-B*13:01, HLA-B*38:02, and HLA-C*08:01 were each carried by a single patient. Similarly, no SA SCAR patients carried any SEA-related HLA-B risk allele, and one patient carried HLA-C*08:01. Conversely, HLA-B*44:03 was uncommon in SEA SCAR patients (1/91) identified by Wang *et al* [6]. Indeed, using multidimensional scaling [39], while US and SA cases and controls demonstrated overlapping HLA class I genotypes, these were distinct from co-trimoxazole-induced SCAR patients among datasets from Japan [40], Thailand [28], and Taiwan [6] (Figure E3). Further, while HLA-A*11:01 was associated with sulfonamide antibiotic-related SCAR in the Japanese population, ‘B*44:03-like’ and ‘B*38:01-like’ cluster alleles were carried by 10/15 patients [40].

HLA class I has evolved under pressure of infectious diseases and although HLA alleles have varied as humans evolved out of Africa, major supertypes are conserved across geographic locations. Analogous to this is the global representation of B44 alleles with SPBS associated with co-trimoxazole-induced SCAR (Figure 4). Intriguingly, HLA-B*13:01 and HLA-B*13:02 evolved separately from a previously shared phylogenetic branch with other ‘B*44:03-like’ alleles including HLA-B*44:03, HLA-B*44:02, and HLA-B*18:01 (Figure E4). Although our study is the first to describe a specific association between HLA-B*44:03 and cotrimoxazole-induced SCAR, it fits with some of the earliest HLA studies of SCAR. Indeed, in a case series in 1987, Roujeau *et al* identified an association between sulphonamide antibiotic-induced TEN and the HLA-B12 serotype [41], which includes B44 and B45 antigens. In 1983,“familial” SJS/TEN was documented in identical twins and their grandfather who all carried HLA-B44 and received a sulfonamide antibiotic, sulfisoxazole [42]. In 1997, Schnyder *et al* reported the HLA-B44-restricted presentation of SMX to T-cells from an SJS/TEN patient [43]. In 2002, Nassif *et al* reported HLA-Cw4-restricted SMX-reactive T-cells from an SJS/TEN patient [44], in keeping with HLA-C*04:01-restricted risk reported here. More recently, T-cells from carriers of HLA-B*13:01 with co-trimoxazole-induced SCAR are reported to proliferate with SMX, but not TMP [6], which is supported by our molecular docking and the higher binding affinity of SMX to HLA-B*13:01 and HLA-B*44:03. Moreover, while similar associations with HLA-B*13:01 are reported for hypersensitivity to dapsone [35] and sulfasalazine [45], we also report carriage of ‘B*44:03-like’ alleles by a small US cohort of sulfasalazine-induced SCAR patients. In keeping with these findings, Fukunaga *et al* recently report an association between salazosulfapyridine (sulfasalazine)-SCAR in a Japanese population and HLA-B*39:01 [46], which we show has SPBS with HLA-B*38:01 and HLA-B*38:02. While these data support shared risk across sulfonamide antibiotics, docking suggests that HLA-B*44:03 might not bind to dapsone, potentially explaining incomplete clinical cross-reactivity observed with co-trimoxazole [38].

**Figure 4.**
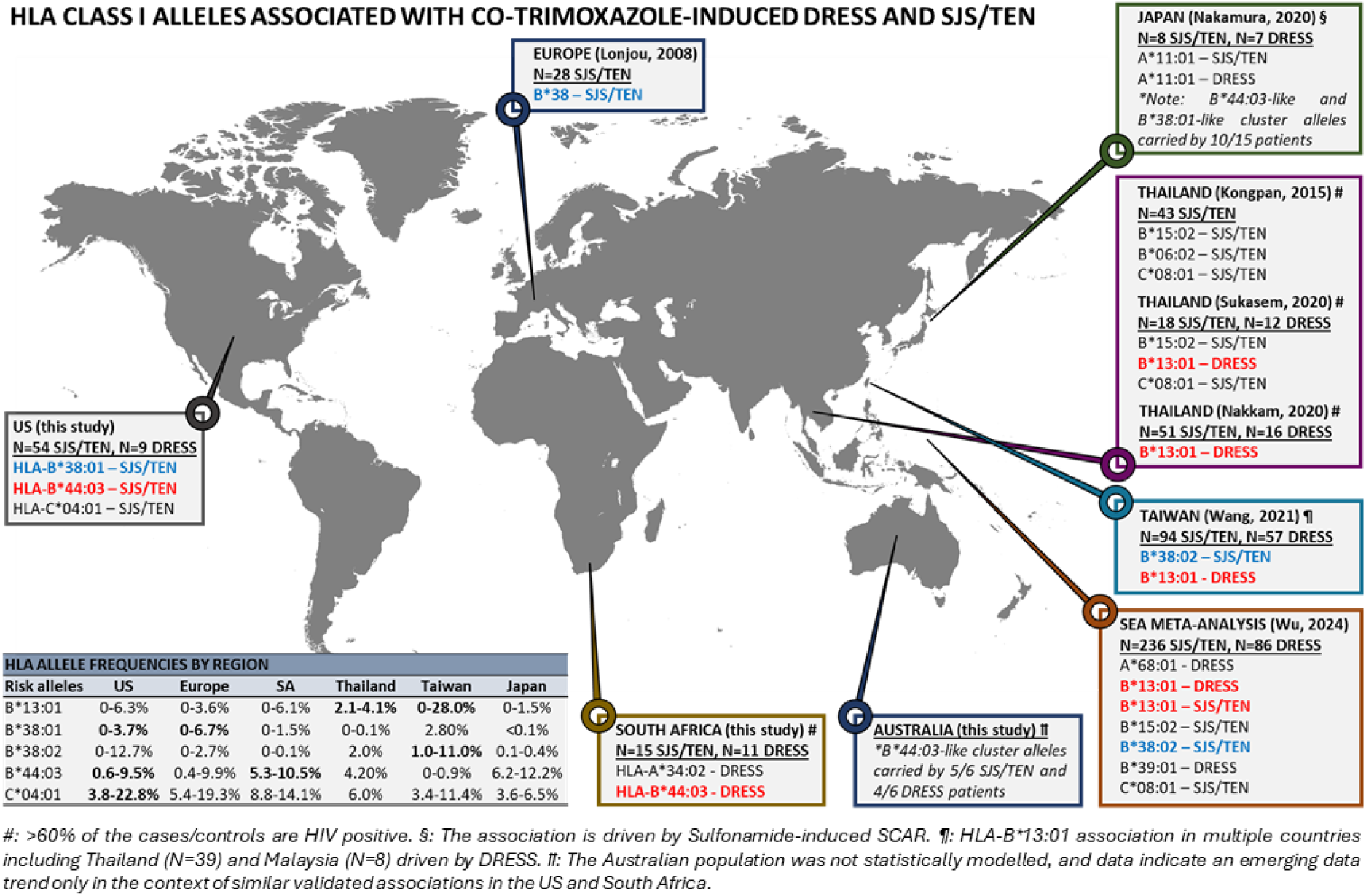
HLA class I risk factors associated with co-trimoxazole induced SCAR in global populations. HLA risk alleles reported in this and previous studies in the US, South Africa, Australia, Europe, Thailand, Taiwan, Malysia, and Japan. Alleles with shared peptide binding specificity across populations are highlighted (red: HLA-B*44:03, HLA-B*13:01; blue: HLA-B*38:01, HLA-B*38:02). Frequencies of risk alleles by region are shown and bolded where specific risk is reported.

While docking and SPBS studies suggest interaction of the drug/metabolite and HLA binding cleft, and both SMX and immunogenic metabolites are shown to stimulate T-cells [32, 43], the HLA-interacting epitope(s) and model by which these activate T-cells remains unknown. However, the original HLA-B44 supertype definition includes B18, B40 (B60 and B61), B44, and B45 serological specificities corresponding to >100 alleles with position 2 specificity for acidic residues and C-terminal specificity for hydrophobic or aromatic residues [23]. These show broad cross-reactivities, with high-affinity binding of 90% of peptides containing B44 super-motifs identified across B44 alleles [22, 23]. This supports the hypothesis that similar risk epitopes, which may be the sentinel epitopes selected within different populations, react across the B44 “cluster”. However, given that only a small number of HLA-predisposed patients will develop co-trimoxazole-induced SCAR, studies to define other risk factors are needed to fully understand risk and immunopathogenesis and make this feasible for preventative screening. Nonetheless, the presence of specific risk alleles has immediate utility in diagnosis to provide adjunctive information to identify the most likely culprit drug, expediting time to effective intervention and guiding future drug safety.

Although limited by small numbers and without complete ascertainment of cases across populations, our study is compelling in its suggestion that HLA risk alleles differ according to specific populations and that these risk alleles have SPBS that will help define specific mechanisms of co-trimoxazole and other sulfonamide antibiotic-induced SCAR. Moreover, as we recently report T-cell receptors (TCR) expressed by pathogenic CD8+ T-cells at the site of tissue damage during co-trimoxazole-SJS/TEN [47], knowledge of both risk TCRαβ and now related HLA risk alleles will enable functional studies to define HLA-TCR-restricted drug epitope(s). Functional studies will also be critical to confirm the functional relevance of SPBS between HLA alleles. Although this may be important for some drugs and not others. However, it is important to note that HLA-B*44:03 is also previously reported as a risk factor for cold medicine (CM)-related SJS/TEN in Indian, Brazilian, and Japanese populations [48, 49], and lamotrigine-induced SJS/TEN in Korea [50] and maculopapular exanthema (MPE) in Thailand [51], albeit in small study cohorts. In the latter study, where HLA-B*44:03 is carried by just 3/10 MPE patients, one other patient expressed HLA-B*13:01 and another expressed HLA-B*40:01; both with SPBS to HLA-B*44:03. In addition, we have previously reported that HLA-C alleles with SPBS define global risk for cutaneous nevirapine hypersensitivity across ancestral groups [52]. Together these data suggest SPBS risk clusters may predispose global population risk for other drugs that may provide important clues to mechanisms and warrant future study.

Study limitations include the smaller number of SA case and population controls, which may have underpowered detection of HLA-C*04:01 risk carried by 23% of SA controls but 46% of co-trimoxazole-induced DRESS patients. Second, unlike the US, where an absence of PLWH among patients aligns with low HIV US population prevalence (<0.5%) [53], the HIV prevalence in SA SCAR patients was 81% but is unknown and certainly lower among pooled population controls. Indeed, while one control database was of healthy individuals [17, 18], the second did not report HIV prevalence [16]. Thus, although shared HLA-B*44:03 risk was identified in both the US and SA, as HLA alleles are associated with HIV control, HIV-matched studies are needed to discern genetic risk for PLWH.

In conclusion, we show that HLA class I alleles with SPBS including HLA-B*44:03 and HLA-B*38:01 but also HLA-C*04:01 predispose co-trimoxazole-induced-SCAR in the US with HLA-B*44:03 generalizing as a DRESS risk factor in SA. Although an HLA allele has the same implications for risk regardless of the population, the primary population risk allele may not be the same, and population-based studies with SPBS and functional tools will more completely define drug-related risk factors that can be used for risk stratification and diagnosis.

## Data Availability

All data produced in the present work are contained in the manuscript

## Acknowledgements

We acknowledge the participation of all DRESS and SJS patients and survivors and the SJS and DRESS syndrome Foundations for their contributions in raising community awareness for the study. We acknowledge John Mings, Blake Stuck, and the VUMC Marketing team for their expertise and guidance in our advertising campaigns for the SJS survivor study. We also gratefully acknowledge the significant expertise of the Immunogenomics and microbial genetics single-cell technologies (IMGSCT) core at VUMC for DNA extraction, and the core genomics laboratories at Murdoch University for sample sequencing, and researchers and clinicians from the Immune-Mediated Adverse Drug Reactions in African HIV Endemic Setting (IMARI-SA) study and African Registry of Severe Cutaneous Adverse Reactions (AFRiSCAR) for the provision of clinically curated SJS/TEN and DRESS patient samples from South Africa. We also acknowledge clinicians and researchers from the Australasian Registry for Severe Cutaneous Adverse Reaction (AUS-SCAR) for their assistance in curating SJS/TEN and DRESS patient samples from Australasia, including but not limited to Amy Legg, Johannes Kern, Abby Douglas, and Ar Kar Aung.

## Abbreviations

HLA: human leukocyte antigen
SCAR: severe cutaneous adverse reaction
SJS/TEN: Stevens-Johnson syndrome and toxic epidermal necrolysis
DRESS: drug reaction with eosinophilia and systemic symptoms
HIV: human immunodeficiency virus
PLWH: persons living with HIV
SEA: Southeast Asian
US: United States
SA: South African
SPBS: shared peptide binding specificity
VUMC: Vanderbilt University Medical Centre
MEEI: Massachusetts Eye and Ear Institute
ALDEN: algorithm for drug causality in epidermal necrolysis
HREC: human research ethics committee
DNA: Deoxyribonucleic acid
EDTA: Ethylenediaminetetraacetic acid
ASHI: American society for Histocompatibility and Immunogenetics
NATA: National Association of Testing Authorities
bp: base pairs
PDB: protein data bank
SMX: sulfamethoxazole
SMX-NO: nitroso-sulfamethoxazole
TMP: trimethoprim
OR: odds ratio
Pc: corrected P value
PPV: positive predictive value
NPV: negative predictive value
NNT: number needed to test.

---

**Figure E1.**
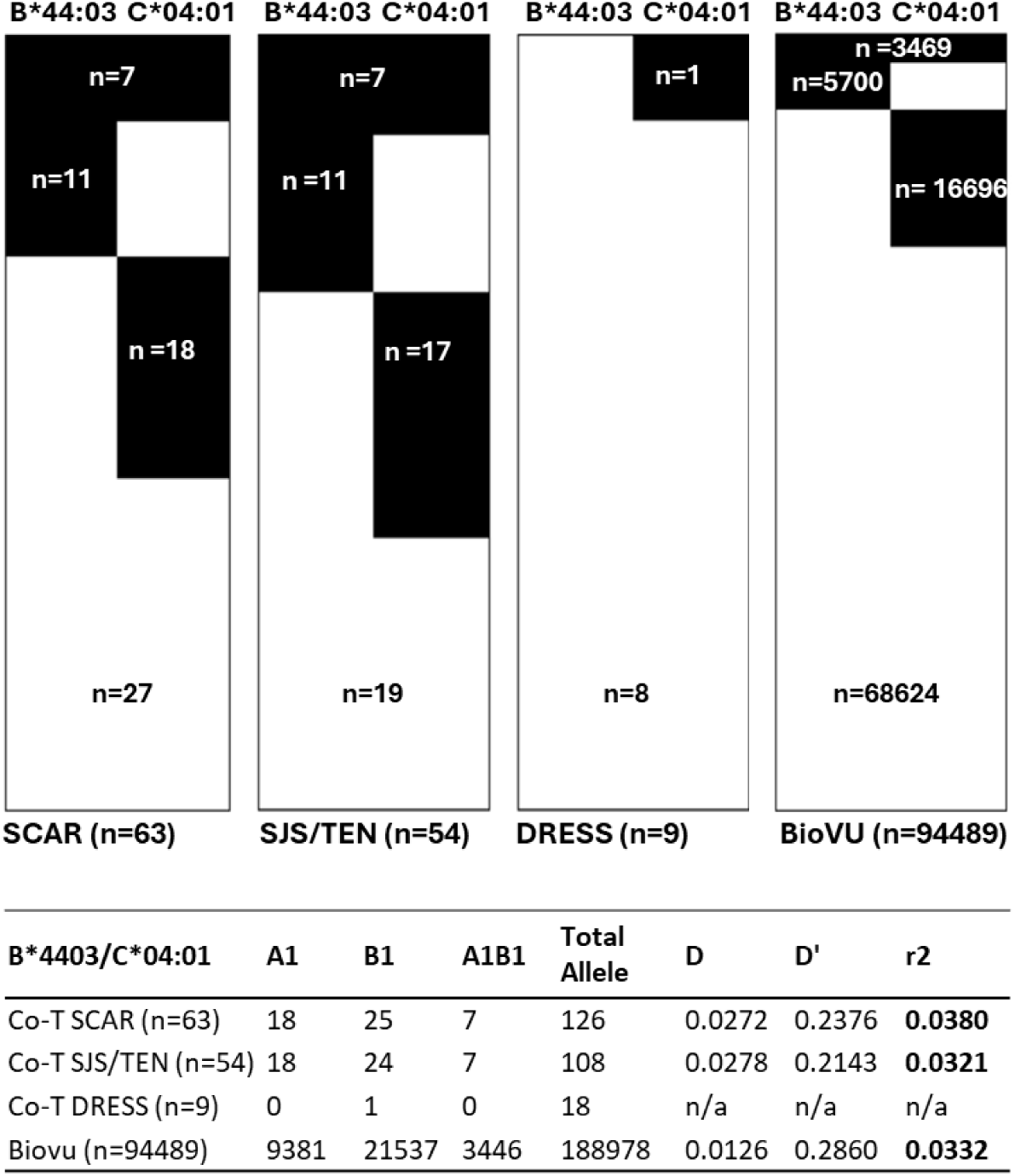
HLA-B*44:03 and HLA-C*04:01 are not in haplotypic association. **(A)** Proportional representation of individuals carrying HLA-B*44:03 and/or HLA-C*04:01 (black) compared to those without (white). **(B)** Haplotype analyses for SCAR patients in the US and BioVU population controls expressed as normalized linkage disequilibrium (D^I^) and correlation coefficient (r^2^) which were both ≈0.

**Figure E2.**
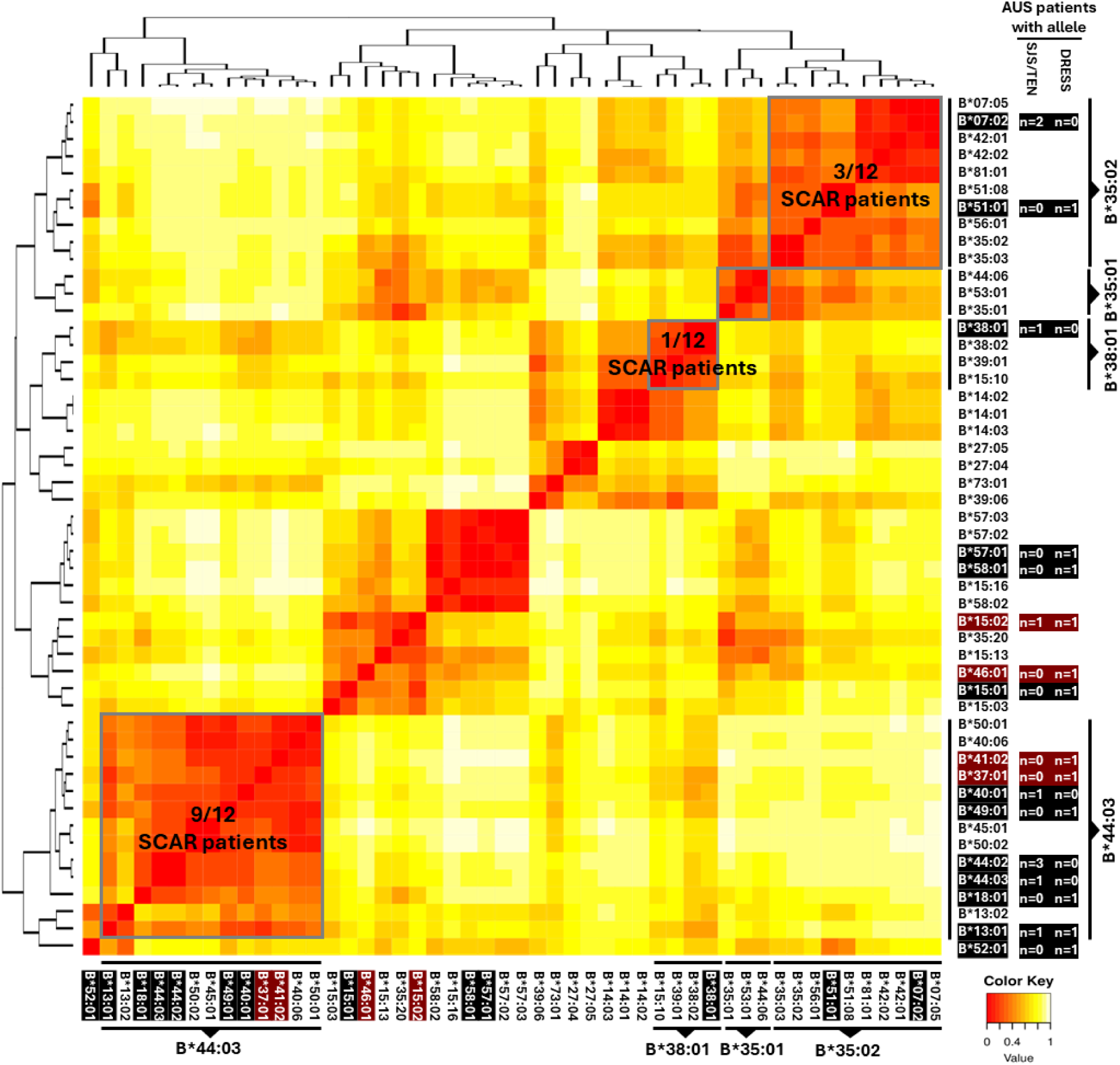
Integration of HLA-B alleles with shared peptide binding specificity in co-trimoxazole-SCAR patients from Australia with those in the US and South Africa. HLA-B alleles are clustered by shared peptide binding (high to low, red to yellow). HLA-B alleles identified in the Australian population and the US or South African population are highlighted black, and those unique to the Australian cohort highlighted red.

**Figure E3.**
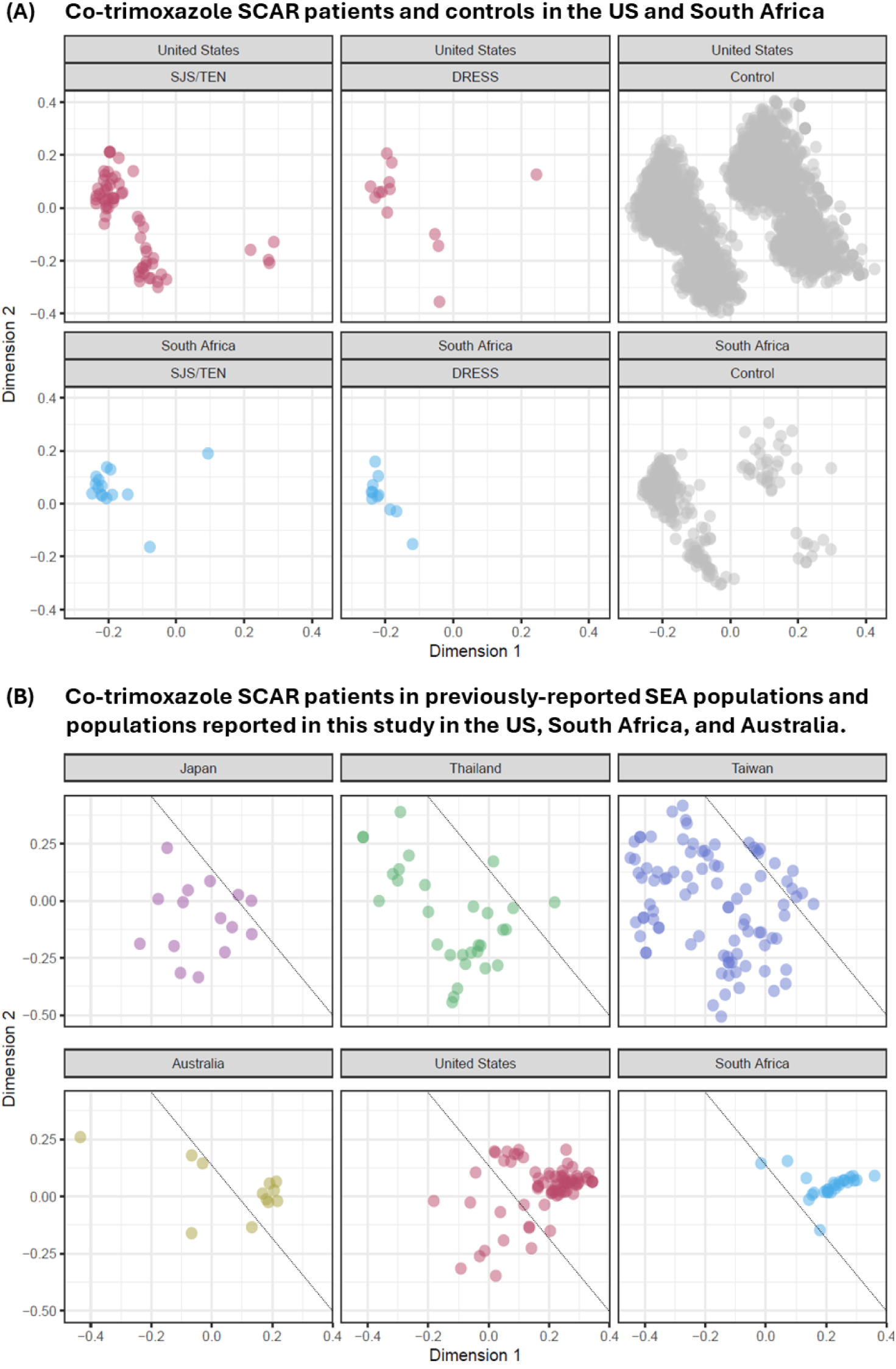
HLA risk alleles for co-trimoxazole-induced SCAR are distinct between Southeast Asian and other global populations. HLA class I genotypes are clustered using pairwise comparisons and Jaccard similarity to generate a similarity matrix for multidimensional comparison of co-trimoxazole-induced SCAR patients from the US and South Africa with **(A)** controls and **(B)** co-trimoxazole-SCAR patients in published datasets from Japan, Thailand, and Taiwan. To ensure balanced allelic representation, the large US control was down sampled to 1,000 subjects per ancestry group.

**Figure E4.**
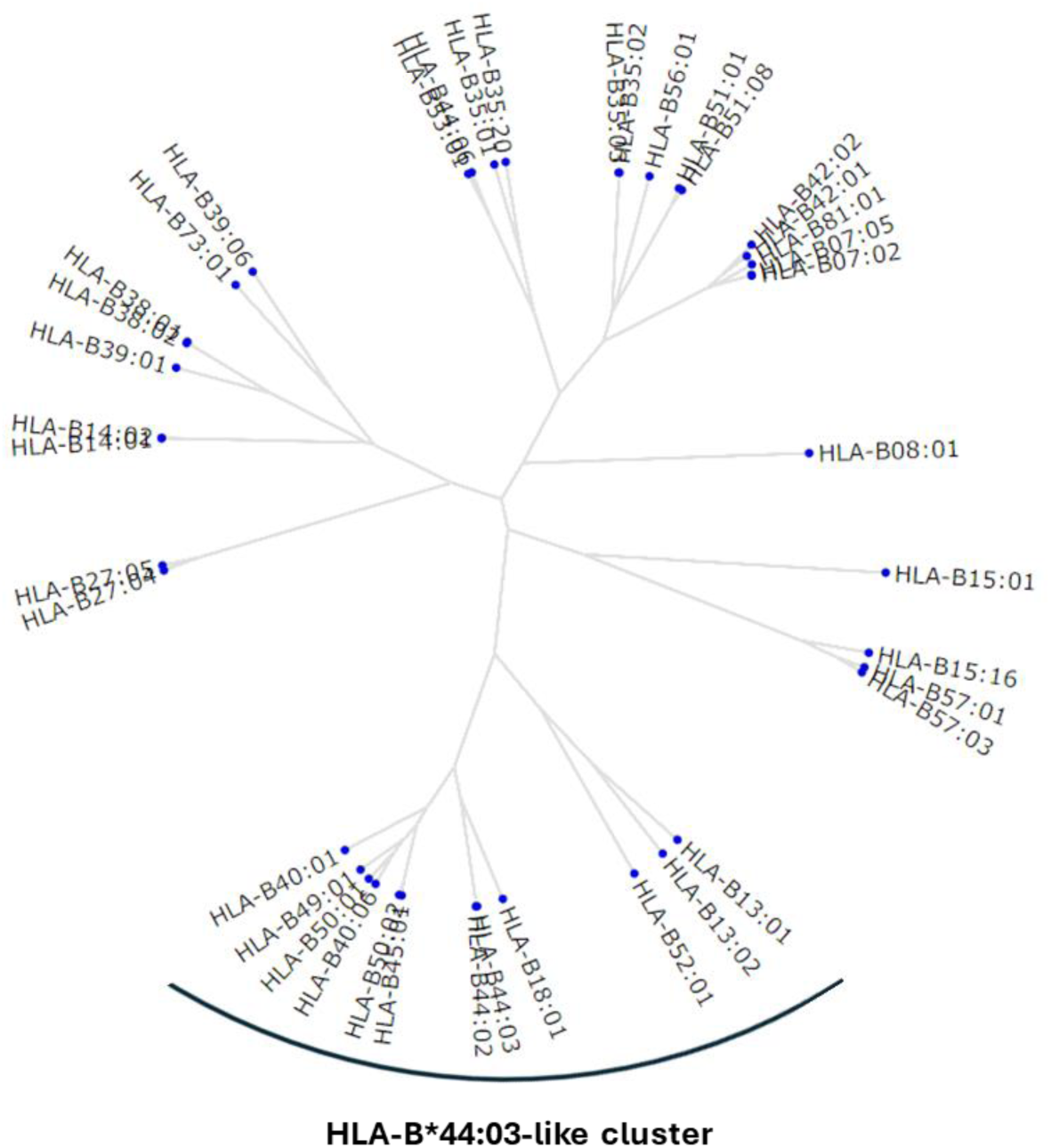
Evolutionary divergence of HLA-B alleles carried by cotrimoxazole-induced SCAR patients in the US. Shared and distinct evolutionary trajectory is shown as a phylogenetic tree generated using MHC cluster 2.0 and the 41 HLA-B alleles identified across all co-trimoxazole-induced SCAR patients in the US cohort. *HLA, human leukocyte antigen*.

**Table E1.** Demographic and clinical characteristics of patients with co-trimoxazole-induced DRESS and SJS/TEN in the US and South Africa. Mean latency (drug initiation to symptom onset) shown in days ± standard deviation. *BSA, body surface area detached; ALDEN, algorithm for drug causality in epidermal necrolysis; HIV, human immunodeficiency virus; N/A, not applicable*.

| Geographic region | US |  | South Africa |  |
| --- | --- | --- | --- | --- |
| Culprit drug | Co-Trimoxazole |  | Co-Trimoxazole |  |
| Phenotype | SJS/TEN | DRESS | SJS/TEN | DRESS |
| Count | N=54 | N=9 | N=15 | N=11 |
| Sex |  |  |  |  |
| Female | 40 (74%) | 8 (89%) | 5 (33%) | 8 (73%) |
| Male | 14 (26%) | 1 (11%) | 10 (67%) | 3 (27%) |
| Skin detachment (%) |  |  |  |  |
| BSA<10% | 23 (43%) | N/A | 0 | N/A |
| BSA>10% | 31 (57%) | N/A | 15 (100%) | N/A |
| Severity |  |  |  |  |
| Latency (days) | 9±6 | 10±7 | 21±27 | 47±55 |
| ALDEN score | 6±1 | N/A | 6±1 | N/A |
| Naranjo Score | 5±1 | 6±1 | 7±1 | 7±1 |
| Regi SCAR | NA | 5±1 | N/A | 5±1 |
| Mortality |  |  |  |  |
| Surviving | 54 (100%) | 9 (100%) | 15 (100%) | 9 (82%) |
| Deceased | 0 | 0 | 0 | 2 (18%) |
| HIV status |  |  |  |  |
| Positive | 0 | 0 | 11 (73%) | 10 (91%) |
| Negative | 54 (100%) | 9 (100%) | 4 (27%) | 1 (9%) |

**Table E2.** HLA class I genotypes of co-trimoxazole- or sulfasalazine-induced SCAR patients in the US. HLA alleles highlighted according to alignments with shared peptide binding specificities (B*44:03-like, green; B*38:01-like, blue) or carriage of HLA-C*04:01 (orange). Ѱ represents patients from the SJS Survivor study. *F, female; M, male*.

| Patient | Culprit drug | Sex | HLA-A allele 1 | HLA-A allele 2 | HLA-B allele 1 | HLA-B allele 2 | HLA-C allele 1 | HLA-C allele 2 | B*44:03-like cluster | B*38:01-like cluster | C*04:01 allele |
| --- | --- | --- | --- | --- | --- | --- | --- | --- | --- | --- | --- |
| US SJS/TEN 1 | Co-trimoxazole | M | A*02:01 | A*02:01 | B*44:02 | B*44:03 | C*05:01 | C*16:01 |  |  |  |
| $\Psi$ US SJS/TEN 2 | Co-trimoxazole | F | A*03:01 | A*30:01 | B*18:01 | B*51:01 | C*01:02 | C*12:03 | | | |
| $\Psi$ US SJS/TEN 3 | Co-trimoxazole | F | A*25:01 | A*68:01 | B*18:01 | B*44:03 | C*12:03 | C*16:01 | | | |
| $\Psi$ US SJS/TEN 4 | Co-trimoxazole | F | A*02:01 | A*29:02 | B*27:05 | B*44:03 | C*02:02 | C*16:01 | | | |
| US SJS/TEN 5 | Co-trimoxazole | F | A*02:01 | A*02:01 | B*44:02 | B*44:06 | C*02:02 | C*05:01 |  |  |  |
| US SJS/TEN 6 | Co-trimoxazole | F | A*02:01 | A*30:01 | B*13:02 | B*15:01 | C*03:04 | C*06:02 |  |  |  |
| US SJS/TEN 7 | Co-trimoxazole | F | A*23:01 | A*30:01 | B*42:02 | B*49:01 | C*07:01 | C*17:01 |  |  |  |
| $\Psi$ US SJS/TEN 8 | Co-trimoxazole | F | A*02:01 | A*30:02 | B*07:02 | B*18:01 | C*05:01 | C*07:02 | | | |
| US SJS/TEN 9 | Co-trimoxazole | F | A*01:02 | A*30:02 | B*44:03 | B*49:01 | C*06:02 | C*07:01 |  |  |  |
| US SJS/TEN 10 | Co-trimoxazole | M | A*01:01 | A*68:01 | B*44:03 | B*57:01 | C*06:02 | C*16:01 |  |  |  |
| $\Psi$ US SJS/TEN 11 | Co-trimoxazole | M | A*03:01 | A*25:01 | B*14:02 | B*18:01 | C*08:02 | C*12:03 | | | |
| $\Psi$ US SJS/TEN 12 | Co-trimoxazole | F | A*26:01 | A*29:02 | B*44:03 | B*49:01 | C*07:01 | C*16:01 | | | |
| $\Psi$ US SJS/TEN 13 | Co-trimoxazole | F | A*29:02 | A*30:01 | B*42:01 | B*44:03 | C*16:01 | C*17:01 | | | |
| $\Psi$ US SJS/TEN 14 | Co-trimoxazole | F | A*02:01 | A*24:02 | B*40:06 | B*44:03 | C*07:01 | C*15:02 | | | |
| $\Psi$ US SJS/TEN 15 | Co-trimoxazole | F | A*24:02 | A*68:01 | B*40:06 | B*44:03 | C*07:01 | C*15:02 | | | |
| US SJS/TEN 16 | Co-trimoxazole | F | A*02:01 | A*29:02 | B*40:01 | B*44:03 | C*03:04 | C*16:01 |  |  |  |
| US SJS/TEN 17 | Co-trimoxazole | M | A*02:01 | A*11:01 | B*07:02 | B*44:02 | C*05:01 | C*07:02 |  |  |  |
| $\Psi$ US SJS/TEN 18 | Co-trimoxazole | F | A*02:01 | A*02:01 | B*40:01 | B*44:03 | C*02:02 | C*03:04 | | | |
| US SJS/TEN 19 | Co-trimoxazole | F | A*01:01 | A*30:02 | B*49:01 | B*52:01 | C*07:01 | C*12:02 |  |  |  |
| US SJS/TEN 20 | Co-trimoxazole | F | A*11:01 | A*32:01 | B*39:06 | B*44:02 | C*05:01 | C*07:02 |  |  |  |
| $\Psi$ US SJS/TEN 21 | Co-trimoxazole | M | A*03:01 | A*03:01 | B*18:01 | B*38:01 | C*02:02 | C*12:03 | | | |
| $\Psi$ US SJS/TEN 22 | Co-trimoxazole | F | A*03:01 | A*26:01 | B*44:03 | B*56:01 | C*01:02 | C*04:01 | | | |
| $\Psi$ US SJS/TEN 23 | Co-trimoxazole | F | A*24:02 | A*31:01 | B*15:01 | B*44:03 | C*03:04 | C*04:01 | | | |
| $\Psi$ US SJS/TEN 24 | Co-trimoxazole | F | A*02:01 | A*24:02 | B*35:01 | B*40:01 | C*03:04 | C*04:01 | | | |
| US SJS/TEN 25 | Co-trimoxazole | F | A*02:01 | A*23:01 | B*08:01 | B*50:02 | C*04:01 | C*07:01 |  |  |  |
| $\Psi$ US SJS/TEN 26 | Co-trimoxazole | F | A*23:01 | A*68:01 | B*44:02 | B*44:03 | C*04:01 | C*07:04 | | | |
| US SJS/TEN 27 | Co-trimoxazole | F | A*02:01 | A*31:01 | B*35:20 | B*44:03 | C*04:01 | C*16:01 |  |  |  |
| US SJS/TEN 28 | Co-trimoxazole | M | A*01:01 | A*03:01 | B*35:01 | B*44:02 | C*04:01 | C*05:01 |  |  |  |
| $\Psi$ US SJS/TEN 29 | Co-trimoxazole | F | A*26:01 | A*33:01 | B*35:03 | B*45:01 | C*04:01 | C*06:02 | | | |
| $\Psi$ US SJS/TEN 30 | Co-trimoxazole | M | A*02:01 | A*24:02 | B*35:02 | B*44:02 | C*04:01 | C*07:04 | | | |
| US SJS/TEN 31 | Co-trimoxazole | F | A*01:01 | A*02:01 | B*27:05 | B*44:03 | C*01:02 | C*04:01 |  |  |  |
| $\Psi$ US SJS/TEN 32 | Co-trimoxazole | F | A*29:02 | A*31:01 | B*40:01 | B*44:03 | C*03:04 | C*04:01 | | | |
| $\Psi$ US SJS/TEN 33 | Co-trimoxazole | F | A*24:02 | A*26:01 | B*35:02 | B*49:01 | C*04:01 | C*07:01 | | | |
| $\Psi$ US SJS/TEN 34 | Co-trimoxazole | M | A*30:02 | A*66:02 | B*44:03 | B*57:03 | C*04:01 | C*07:01 | | | |
| US SJS/TEN 35 | Co-trimoxazole | F | A*02:01 | A*03:01 | B*40:01 | B*53:01 | C*03:04 | C*04:01 |  |  |  |
| US SJS/TEN 36 | Co-trimoxazole | M | A*01:01 | A*03:01 | B*08:01 | B*35:01 | C*04:01 | C*07:01 |  |  |  |
| US SJS/TEN 37 | Co-trimoxazole | F | A*02:02 | A*36:01 | B*15:16 | B*53:01 | C*04:01 | C*14:02 |  |  |  |
| $\Psi$ US SJS/TEN 38 | Co-trimoxazole | M | A*03:01 | A*11:01 | B*15:01 | B*35:01 | C*03:03 | C*04:01 | | | |
| $\Psi$ US SJS/TEN 39 | Co-trimoxazole | F | A*02:01 | A*11:01 | B*15:01 | B*35:01 | C*03:04 | C*04:01 | | | |
| $\Psi$ US SJS/TEN 40 | Co-trimoxazole | F | A*33:03 | A*68:02 | B*53:01 | B*53:01 | C*04:01 | C*04:01 | | | |
| $\Psi$ US SJS/TEN 41 | Co-trimoxazole | M | A*01:01 | A*03:01 | B*08:01 | B*35:01 | C*04:01 | C*07:01 | | | |
| $\Psi$ US SJS/TEN 42 | Co-trimoxazole | F | A*02:01 | A*24:02 | B*27:05 | B*35:02 | C*02:02 | C*04:01 | | | |
| $\Psi$ US SJS/TEN 43 | Co-trimoxazole | F | A*03:01 | A*68:02 | B*35:01 | B*81:01 | C*04:01 | C*18:01 | | | |
| US SJS/TEN 44 | Co-trimoxazole | F | A*03:01 | A*30:01 | B*35:01 | B*42:01 | C*04:01 | C*17:01 |  |  |  |
| US SJS/TEN 45 | Co-trimoxazole | F | A*01:01 | A*03:01 | B*08:01 | B*35:01 | C*04:01 | C*07:01 |  |  |  |
| $\Psi$ US SJS/TEN 46 | Co-trimoxazole | M | A*02:01 | A*02:30 | B*38:01 | B*38:01 | C*12:03 | C*12:03 | | | |
| $\Psi$ US SJS/TEN 47 | Co-trimoxazole | F | A*24:02 | A*29:02 | B*14:02 | B*38:01 | C*08:02 | C*12:03 | | | |
| US SJS/TEN 48 | Co-trimoxazole | F | A*02:03 | A*29:01 | B*07:05 | B*38:02 | C*07:02 | C*15:05 |  |  |  |
| US SJS/TEN 49 | Co-trimoxazole | M | A*11:01 | A*31:01 | B*27:04 | B*39:01 | C*07:02 | C*07:02 |  |  |  |
| $\Psi$ US SJS/TEN 50 | Co-trimoxazole | F | A*26:01 | A*30:01 | B*38:01 | B*42:01 | C*12:03 | C*17:01 | | | |
| US SJS/TEN 51 | Co-trimoxazole | F | A*02:01 | A*11:01 | B*38:01 | B*51:01 | C*12:03 | C*15:02 |  |  |  |
| $\Psi$ US SJS/TEN 52 | Co-trimoxazole | F | A*01:01 | A*30:01 | B*08:01 | B*08:01 | C*07:01 | C*07:02 | | | |
| $\Psi$ US SJS/TEN 53 | Co-trimoxazole | F | A*02:01 | A*32:01 | B*42:02 | B*73:01 | C*15:05 | C*17:01 | | | |
| US SJS/TEN 54 | Co-trimoxazole | M | A*02:01 | A*03:01 | B*07:02 | B*51:08 | C*07:02 | C*16:02 |  |  |  |
| $\Psi$ US SJS/TEN 55 | Sulfasalazine | F | A*03:01 | A*29:02 | B*08:01 | B*44:03 | C*07:01 | C*16:01 | | | |
| $\Psi$ US SJS/TEN 56 | Sulfasalazine | F | A*02:01 | A*29:02 | B*44:02 | B*44:03 | C*05:01 | C*16:01 | | | |
| US SJS/TEN 57 | Sulfasalazine | F | A*03:01 | A*03:01 | B*15:01 | B*35:01 | C*03:03 | C*04:01 |  |  |  |
| $\Psi$ US SJS/TEN 58 | Sulfasalazine | M | A*02:01 | A*11:01 | B*07:02 | B*35:01 | C*04:01 | C*07:02 | | | |
| $\Psi$ US SJS/TEN 59 | Sulfasalazine | F | A*30:02 | A*68:01 | B*38:01 | B*56:01 | C*01:02 | C*12:03 | | | |
| $\Psi$ US SJS/TEN 60 | Sulfasalazine | F | A*01:01 | A*23:01 | B*14:01 | B*38:01 | C*08:02 | C*12:03 | | | |
| $\Psi$ US SJS/TEN 61 | Sulfasalazine | F | A*26:01 | A*74:01 | B*38:01 | B*42:01 | C*12:03 | C*17:01 | | | |
| US DRESS 1 | Co-trimoxazole | M | A*01:01 | A*03:02 | B*08:01 | B*50:01 | C*06:02 | C*07:01 |  |  |  |
| US DRESS 2 | Co-trimoxazole | F | A*02:06 | A*34:02 | B*08:01 | B*40:06 | C*07:01 | C*08:01 |  |  |  |
| US DRESS 3 | Co-trimoxazole | F | A*02:01 | A*03:01 | B*08:01 | B*40:01 | C*03:04 | C*07:01 |  |  |  |
| US DRESS 4 | Co-trimoxazole | F | A*11:01 | A*24:02 | B*13:01 | B*44:02 | C*04:03 | C*16:04 |  |  |  |
| US DRESS 5 | Co-trimoxazole | F | A*25:01 | A*30:02 | B*18:01 | B*50:01 | C*06:02 | C*12:03 |  |  |  |
| US DRESS 6 | Co-trimoxazole | F | A*26:01 | A*01:01 | B*38:01 | B*08:01 | C*12:03 | C*07:01 |  |  |  |
| US DRESS 7 | Co-trimoxazole | F | A*01:01 | A*25:01 | B*35:02 | B*39:01 | C*04:01 | C*12:03 |  |  |  |
| US DRESS 8 | Co-trimoxazole | F | A*03:01 | A*26:01 | B*07:02 | B*07:02 | C*07:02 | C*07:02 |  |  |  |
| US DRESS 9 | Co-trimoxazole | F | A*01:01 | A*02:01 | B*08:01 | B*15:01 | C*03:04 | C*07:01 |  |  |  |
| US DRESS 10 | Sulfasalazine | F | A*02:01 | A*29:02 | B*40:01 | B*51:01 | C*03:04 | C*07:02 |  |  |  |
| US DRESS 11 | Sulfasalazine | F | A*03:01 | A*68:02 | B*44:03 | B*53:01 | C*04:01 | C*04:01 |  |  |  |
| US DRESS 12 | Sulfasalazine | F | A*30:02 | A*68:02 | B*53:01 | B*57:03 | C*04:01 | C*18:01 |  |  |  |
| US DRESS 13 | Sulfasalazine | F | A*33:03 | A*36:01 | B*53:01 | B*53:01 | C*04:01 | C*06:02 |  |  |  |

**Table E3.**
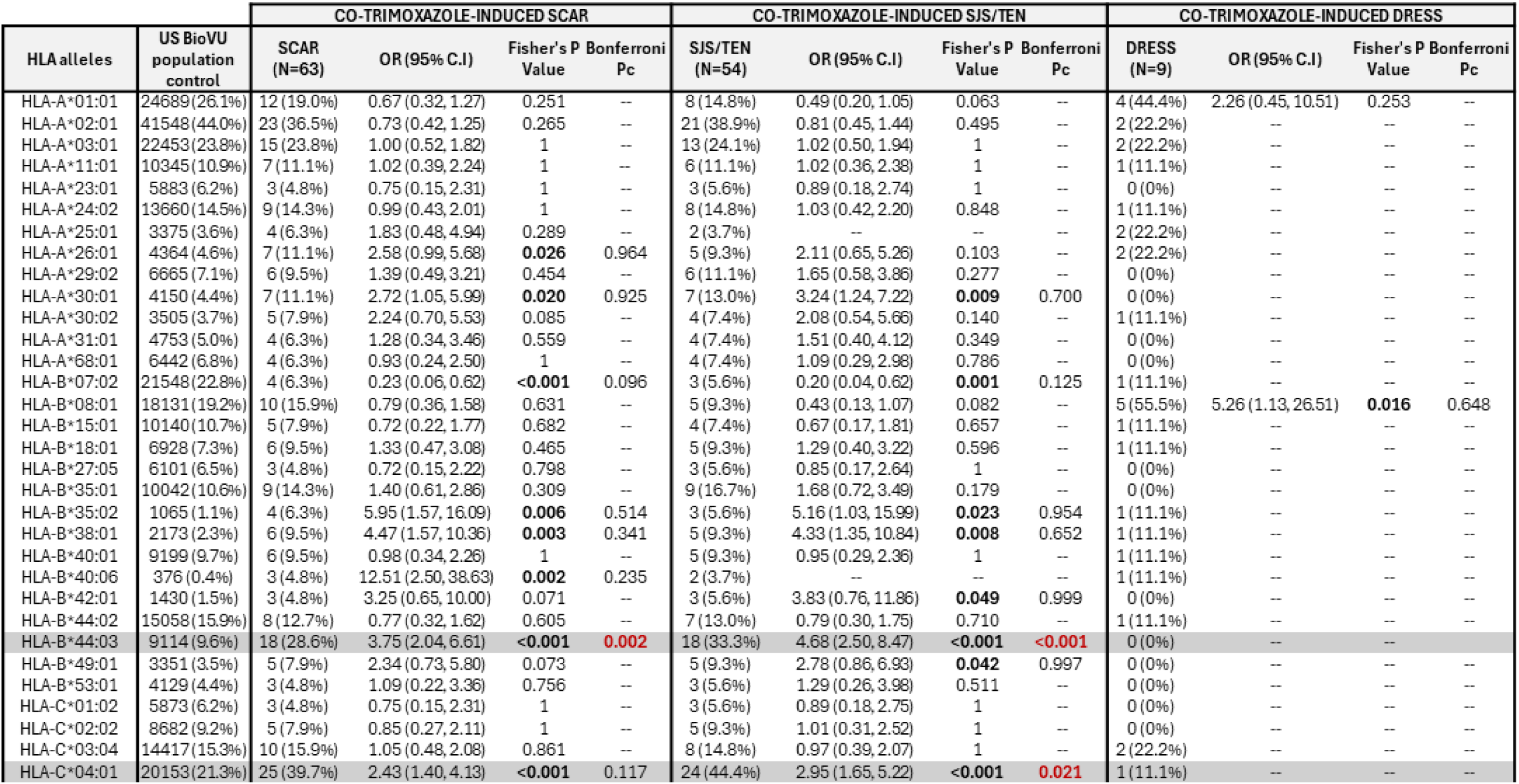
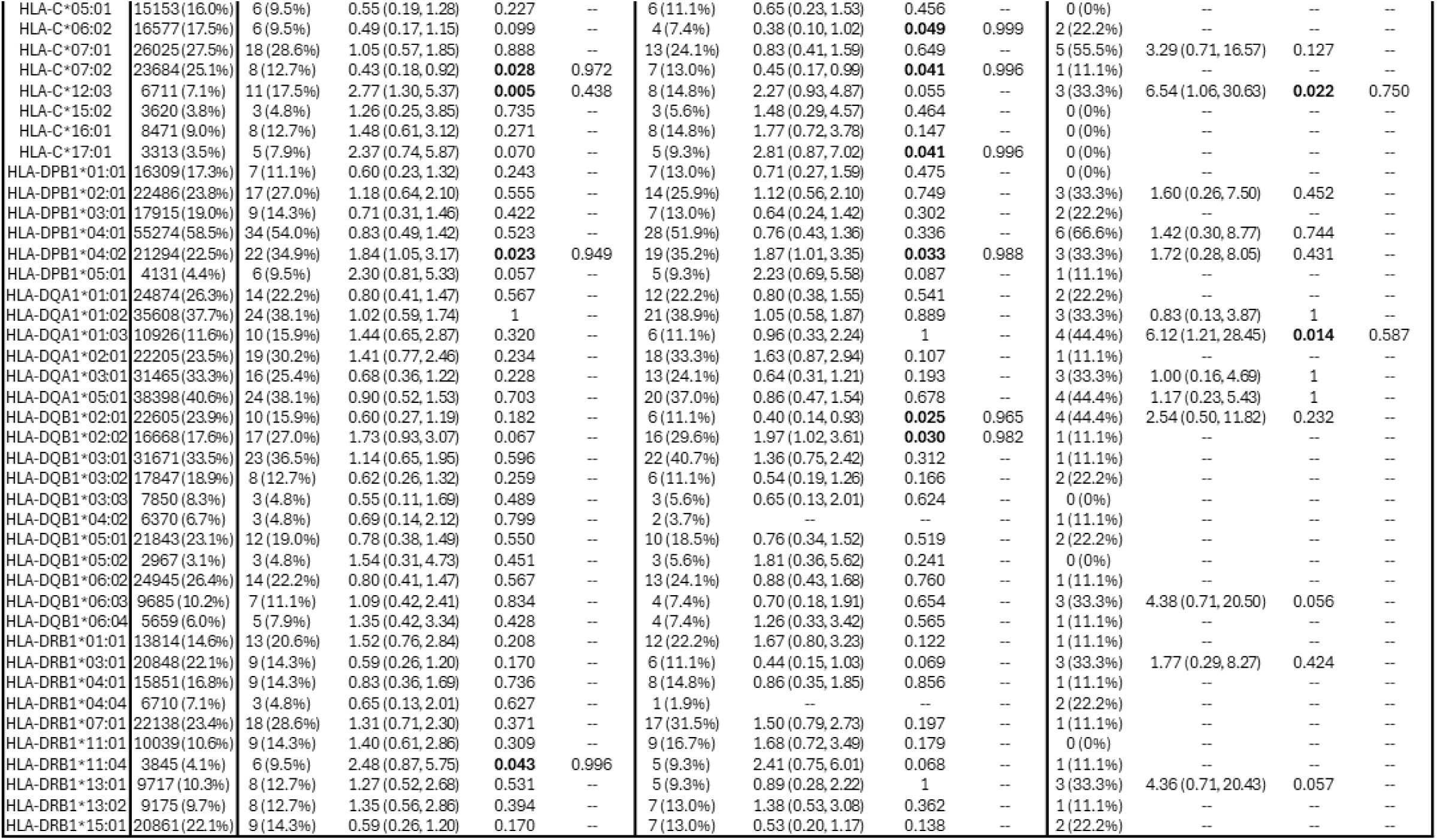
Univariate HLA class I and II association analyses for co-trimoxazole-induced SCAR, SJS/TEN, and DRESS in the US cohort compared to the BioVU population control. P values calculated using Fisher’s exact test with Bonferroni correction. Significant (Pc<0.05) alleles are highlighted grey. HLA alleles carried by ≤2 patients were removed from the table. *OR, odds ratio; CI, confidence interval*.

**Table E4.**
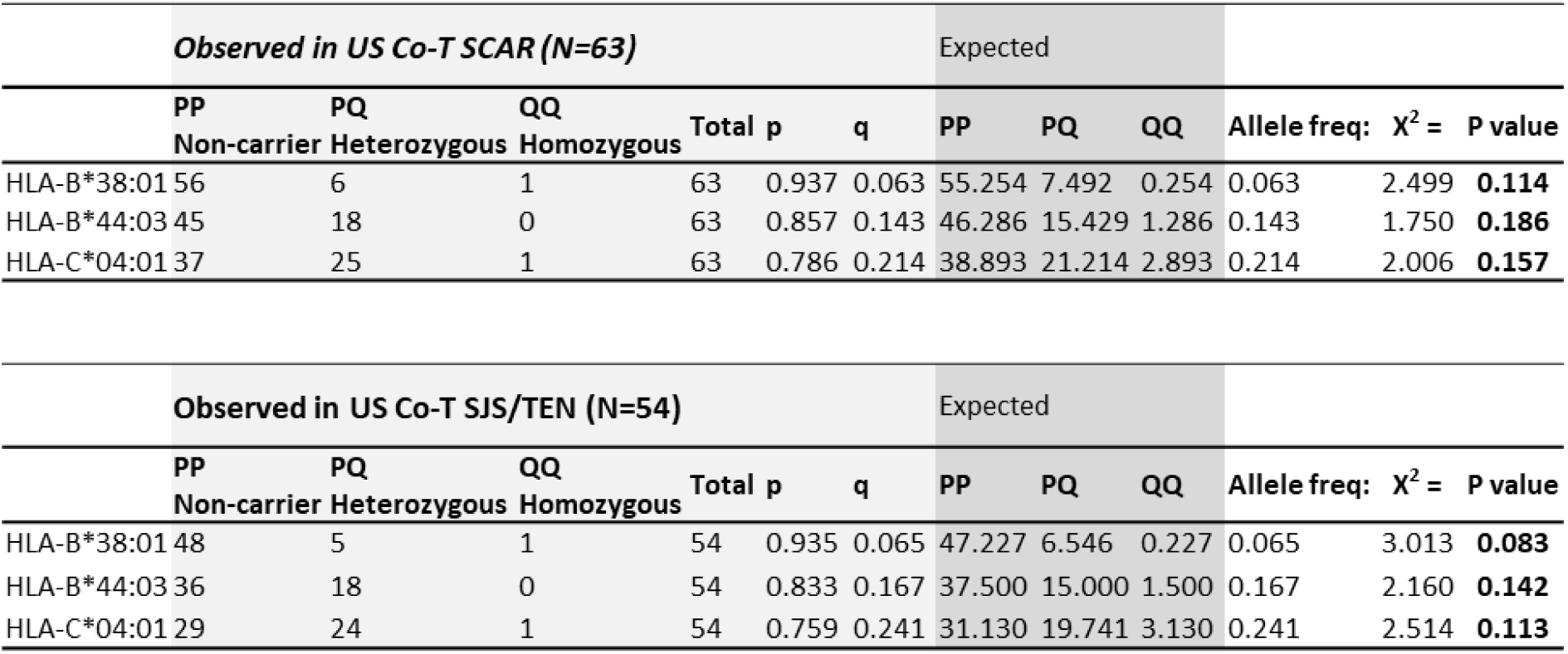
The reported HLA alleles in US SCAR and SJS/TEN patients form Hardy Weinberg equilibrium.

**Table E5.**
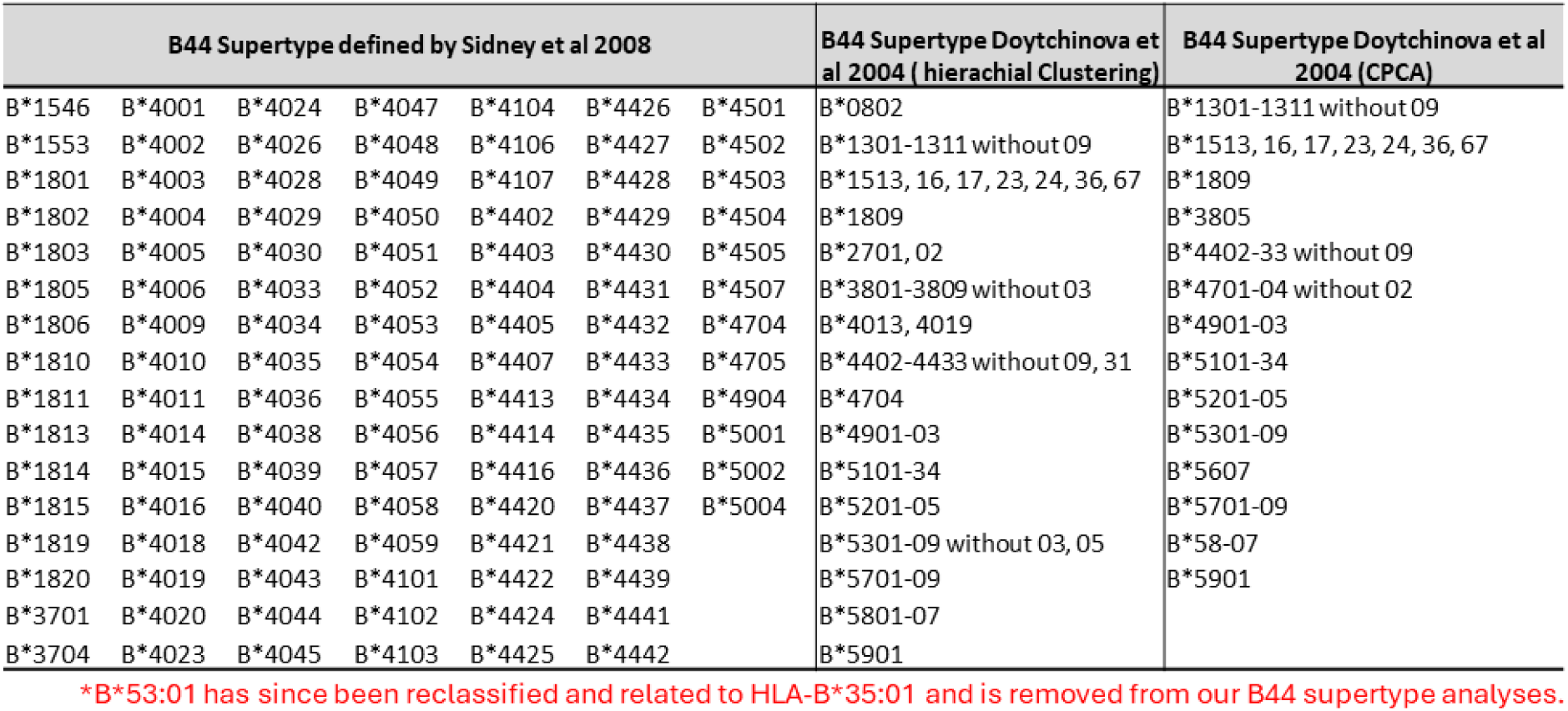
B44 supertype alleles as defined by current in literature. HLA-B alleles are listed which were defined by three different clustering analyses by Sidney in 2003 and 2008 and Doytchinova in 2004 using hierarchical clustering or CPCA. *CPCA, consensus principal component analysis*.

| B44 Supertype defined by Sidney et al 2008 |  |  |  |  |  |  | B44 Supertype Doytchinova et al 2004 ( hierarchical Clustering) | B44 Supertype Doytchinova et al 2004 (CPCA) |
| --- | --- | --- | --- | --- | --- | --- | --- | --- |
| B*1546 | B*4001 | B*4024 | B*4047 | B*4104 | B*4426 | B*4501 | B*0802 | B*1301-1311 without 09 |
| B*1553 | B*4002 | B*4026 | B*4048 | B*4106 | B*4427 | B*4502 | B*1301-1311 without 09 | B*1513, 16, 17, 23, 24, 36, 67 |
| B*1801 | B*4003 | B*4028 | B*4049 | B*4107 | B*4428 | B*4503 | B*1513, 16, 17, 23, 24, 36, 67 | B*1809 |
| B*1802 | B*4004 | B*4029 | B*4050 | B*4402 | B*4429 | B*4504 | B*1809 | B*3805 |
| B*1803 | B*4005 | B*4030 | B*4051 | B*4403 | B*4430 | B*4505 | B*2701, 02 | B*4402-33 without 09 |
| B*1805 | B*4006 | B*4033 | B*4052 | B*4404 | B*4431 | B*4507 | B*3801-3809 without 03 | B*4701-04 without 02 |
| B*1806 | B*4009 | B*4034 | B*4053 | B*4405 | B*4432 | B*4704 | B*4013, 4019 | B*4901-03 |
| B*1810 | B*4010 | B*4035 | B*4054 | B*4407 | B*4433 | B*4705 | B*4402-4433 without 09, 31 | B*5101-34 |
| B*1811 | B*4011 | B*4036 | B*4055 | B*4413 | B*4434 | B*4904 | B*4704 | B*5201-05 |
| B*1813 | B*4014 | B*4038 | B*4056 | B*4414 | B*4435 | B*5001 | B*4901-03 | B*5301-09 |
| B*1814 | B*4015 | B*4039 | B*4057 | B*4416 | B*4436 | B*5002 | B*5101-34 | B*5607 |
| B*1815 | B*4016 | B*4040 | B*4058 | B*4420 | B*4437 | B*5004 | B*5201-05 | B*5701-09 |
| B*1819 | B*4018 | B*4042 | B*4059 | B*4421 | B*4438 |  | B*5301-09 without 03, 05 | B*58-07 |
| B*1820 | B*4019 | B*4043 | B*4101 | B*4422 | B*4439 |  | B*5701-09 | B*5901 |
| B*3701 | B*4020 | B*4044 | B*4102 | B*4424 | B*4441 |  | B*5801-07 |  |
| B*3704 | B*4023 | B*4045 | B*4103 | B*4425 | B*4442 |  | B*5901 |  |

\*B\*53:01 has since been reclassified and related to HLA-B\*35:01 and is removed from our B44 supertype analyses.

**Table E6.** Predictive value of individual and grouped HLA risk alleles in the US population. *PPV, positive predictive value; NPV, negative predictive value; NNT, number needed to test*.

**Co-trimoxazole-SCAR incidence of 0.1%**
| HLA risk | PPV | NPV | Sensitivity | Specificity | NNT | BioVU screened out |
| --- | --- | --- | --- | --- | --- | --- |
| B*38:01 | 0.4% | 99.9% | 9.5% | 97.4% | 9932 | 2% |
| B*44:03 | 0.3% | 99.9% | 28.6% | 89.2% | 3311 | 10% |
| C*04:01 | 0.2% | 99.9% | 39.7% | 76.1% | 2384 | 21% |
| B*38:01, B*44:03, C*04:01 | 0.2% | 99.9% | 65.1% | 67.3% | 1453 | 31% |
| B*44:03-like cluster | 0.1% | 99.9% | 65.1% | 51.2% | 1453 | 42% |
| B*38:01-like cluster | 0.2% | 99.9% | 14.3% | 93.4% | 6621 | 6% |
| B*44:03-like cluster, B*38:01-like cluster | 0.2% | 99.9% | 76.2% | 46.4% | 1241 | 53% |
| B*44:03-like cluster, B*38:01-like cluster, and C*04:01 | 0.1% | 100.0% | 93.7% | 32.7% | 1010 | 63% |

**Table E7.** HLA class I genotypes of co-trimoxazole-induced SCAR patients in South Africa. HLA alleles are highlighted according to alignments with shared peptide binding specificities (B*44:03-like, green; B*38:01-like, blue) or carriage of HLA-C*04:01 (orange). *F, female; M, male*.

| Patient | Culprit drug | Sex | HLA-A<br>allele 1 | HLA-A<br>allele 2 | HLA-B<br>allele 1 | HLA-B<br>allele 2 | HLA-C<br>allele 1 | HLA-C<br>allele 2 | B*44:03-like<br>cluster | B*38:01-like<br>cluster | C*04:01<br>allele |
| --- | --- | --- | --- | --- | --- | --- | --- | --- | --- | --- | --- |
| SA SJS/TEN 1 | Co-trimoxazole | F | A*02:01 | A*24:02 | B*15:13 | B*18:01 | C*07:01 | C*08:01 |  |  |  |
| SA SJS/TEN 2 | Co-trimoxazole | M | A*23:01 | A*68:01 | B*45:01 | B*58:02 | C*06:02 | C*06:02 |  |  |  |
| SA SJS/TEN 3 | Co-trimoxazole | M | A*30:01 | A*74:01 | B*42:01 | B*45:01 | C*06:02 | C*17:01 |  |  |  |
| SA SJS/TEN 4 | Co-trimoxazole | F | A*30:04 | A*68:01 | B*44:03 | B*58:02 | C*02:10 | C*06:02 |  |  |  |
| SA SJS/TEN 5 | Co-trimoxazole | M | A*30:02 | A*30:02 | B*08:01 | B*44:03 | C*04:01 | C*07:01 |  |  |  |
| SA SJS/TEN 6 | Co-trimoxazole | M | A*02:02 | A*32:01 | B*15:16 | B*44:03 | C*04:01 | C*14:02 |  |  |  |
| SA SJS/TEN 7 | Co-trimoxazole | M | A*23:01 | A*23:01 | B*15:10 | B*58:02 | C*06:02 | C*16:01 |  |  |  |
| SA SJS/TEN 8 | Co-trimoxazole | F | A*29:02 | A*30:01 | B*42:01 | B*81:01 | C*04:01 | C*17:01 |  |  |  |
| SA SJS/TEN 9 | Co-trimoxazole | M | A*02:02 | A*74:01 | B*15:03 | B*53:01 | C*02:10 | C*04:01 |  |  |  |
| SA SJS/TEN 10 | Co-trimoxazole | M | A*01:01 | A*66:01 | B*58:02 | B*81:01 | C*06:02 | C*18:01 |  |  |  |
| SA SJS/TEN 11 | Co-trimoxazole | M | A*30:01 | A*30:01 | B*42:01 | B*42:02 | C*17:01 | C*17:01 |  |  |  |
| SA SJS/TEN 12 | Co-trimoxazole | M | A*02:05 | A*03:01 | B*08:01 | B*58:02 | C*06:02 | C*07:02 |  |  |  |
| SA SJS/TEN 13 | Co-trimoxazole | M | A*01:01 | A*32:01 | B*07:02 | B*81:01 | C*01:02 | C*18:01 |  |  |  |
| SA SJS/TEN 14 | Co-trimoxazole | F | A*23:01 | A*30:02 | B*57:03 | B*58:01 | C*07:01 | C*18:01 |  |  |  |
| SA SJS/TEN 15 | Co-trimoxazole | F | A*29:02 | A*74:01 | B*14:03 | B*15:03 | C*02:10 | C*08:02 |  |  |  |
| SA DRESS 1 | Co-trimoxazole | F | A*29:02 | A*34:02 | B*42:01 | B*44:03 | C*07:01 | C*17:01 |  |  |  |
| SA DRESS 2 | Co-trimoxazole | F | A*33:03 | A*68:01 | B*44:03 | B*58:02 | C*06:02 | C*07:01 |  |  |  |
| SA DRESS 3 | Co-trimoxazole | F | A*02:01 | A*68:01 | B*13:02 | B*42:01 | C*06:02 | C*17:01 |  |  |  |
| SA DRESS 4 | Co-trimoxazole | F | A*11:01 | A*34:02 | B*35:01 | B*44:03 | C*04:01 | C*04:01 |  |  |  |
| SA DRESS 5 | Co-trimoxazole | M | A*34:02 | A*68:02 | B*44:03 | B*57:02 | C*04:01 | C*18:01 |  |  |  |
| SA DRESS 6 | Co-trimoxazole | F | A*34:02 | A*68:01 | B*27:05 | B*44:03 | C*02:02 | C*04:01 |  |  |  |
| SA DRESS 7 | Co-trimoxazole | F | A*32:01 | A*68:02 | B*15:10 | B*44:03 | C*02:10 | C*03:04 |  |  |  |
| SA DRESS 8 | Co-trimoxazole | M | A*68:02 | A*68:02 | B*15:10 | B*15:16 | C*03:04 | C*03:04 |  |  |  |
| SA DRESS 9 | Co-trimoxazole | M | A*33:01 | A*33:03 | B*42:01 | B*53:01 | C*04:01 | C*17:01 |  |  |  |
| SA DRESS 10 | Co-trimoxazole | F | A*23:01 | A*68:02 | B*15:03 | B*53:01 | C*02:10 | C*04:01 |  |  |  |
| SA DRESS 11 | Co-trimoxazole | F | A*02:02 | A*29:02 | B*42:01 | B*58:02 | C*06:02 | C*17:01 |  |  |  |

**Table E8.** Univariate HLA class I association analyses for co-trimoxazole-induced SCAR, SJS/TEN, and DRESS in the South African cohort compared to the population control (n=351). P values calculated using Fisher’s exact test with Bonferroni correction. Significant (Pc<0.05) alleles are highlighted grey. HLA alleles carried by ≤2 patients were removed from the table. *OR, odds ratio; CI, confidence interval*.

| HLA alleles | SA population control | CO-TRIMOXAZOLE-INDUCED SCAR |  |  |  | CO-TRIMOXAZOLE-INDUCED SJS/TEN |  |  |  | CO-TRIMOXAZOLE-INDUCED DRESS |  |  |  |
| --- | --- | --- | --- | --- | --- | --- | --- | --- | --- | --- | --- | --- | --- |
|  |  | SCAR (N=26) | OR (95% C.I.) | Fisher's P Value | Bonferroni Pc | SJS/TEN (N=15) | OR (95% C.I.) | Fisher's P Value | Bonferroni Pc | DRESS (N=11) | OR (95% C.I.) | Fisher's P Value | Bonferroni Pc |
| HLA-A*02:02 | 11 (3.1%) | 3 (11.5%) | 4.01 (17.52, 13.34) | 0.064 | -- | 2 (13.3%) | -- | -- | -- | 1 (9.1%) | -- | -- | -- |
| HLA-A*23:01 | 40 (11.4%) | 4 (15.4%) | 1.41 (41.39, 12.31) | 0.526 | -- | 3 (20.0%) | 1.94 (0.34, 7.62) | 0.400 | -- | 1 (9.1%) | -- | -- | -- |
| HLA-A*29:02 | 31 (8.8%) | 4 (15.4%) | 1.87 (13.53, 13.32) | 0.285 | -- | 2 (13.3%) | -- | -- | -- | 2 (18.2%) | -- | -- | -- |
| HLA-A*30:01 | 47 (13.4%) | 3 (11.5%) | 0.84 (19.65, 12.30) | 1 | -- | 3 (20.0%) | 1.61 (0.28, 6.30) | 0.442 | -- | 0 (0.0%) | -- | -- | -- |
| HLA-A*32:01 | 11 (3.1%) | 3 (11.5%) | 4.01 (18., 13.34) | 0.064 | -- | 2 (13.3%) | -- | -- | -- | 1 (9.1%) | -- | -- | -- |
| HLA-A*34:02 | 17 (4.8%) | 4 (15.4%) | 3.55 (2.91, 15.33) | **0.048** | 0.931 | 0 (0%) | -- | -- | -- | 4 (36.4%) | 11.04 (2.16, 48.72) | **0.002** | **0.086** |
| HLA-A*68:01 | 20 (5.7%) | 5 (19.2%) | 3.92 (4.35, 13.33) | **0.021** | 0.696 | 2 (13.3%) | -- | -- | -- | 3 (27.3%) | 6.14 (0.98, 28.26) | **0.027** | **0.676** |
| HLA-A*68:02 | 45 (12.8%) | 4 (15.4%) | 1.24 (6.27, 15.30) | 0.761 | -- | 0 (0.0%) | -- | -- | -- | 4 (36.4%) | 3.86 (0.80, 15.93) | **0.048** | **0.871** |
| HLA-A*74:01 | 19 (5.4%) | 3 (11.5%) |  | 0.187 | -- | 3 (20.0%) | 4.34 (0.73, 18.08) | 0.054 | -- | 0 (0.0%) | -- | -- | -- |
| HLA-B*15:03 | 42 (12.0%) | 3 (11.5%) |  | 1 | -- | 2 (13.3%) | -- | -- | -- | 1 (9.1%) | -- | -- | -- |
| HLA-B*15:10 | 44 (12.5%) | 3 (11.5%) | 0.91 (35.88, 14.30) | 1 | -- | 1 (6.7%) | -- | -- | -- | 2 (18.2%) | -- | -- | -- |
| HLA-B*42:01 | 33 (9.4%) | 7 (26.9%) | 3.53 (3., 12.31) | **0.013** | 0.506 | 3 (20.0%) | 2.40 (0.41, 9.54) | 0.174 | -- | 4 (36.4%) | 5.46 (1.11, 22.84) | **0.018** | **0.537** |
| HLA-B*44:03 | 35 (10.0%) | 9 (34.6%) | 4.75 (1.73, 12.31) | **0.001** | 0.066 | 3 (20.0%) | 2.25 (0.39, 8.91) | 0.196 | -- | 6 (54.5%) | 10.69 (2.57, 46.75) | **<0.001** | **0.019** |
| HLA-B*53:01 | 24 (6.8%) | 3 (11.5%) | 1.77 (11.34, 14.32) | 0.417 | -- | 1 (6.7%) | -- | -- | -- | 2 (18.2%) | -- | -- | -- |
| HLA-B*58:02 | 64 (18.2%) | 7 (26.9%) | 1.65 (38.26, 10.28) | 0.298 | -- | 5 (33.3%) | 2.24 (0.58, 7.48) | 0.172 | -- | 2 (18.2%) | -- | -- | -- |
| HLA-B*81:01 | 17 (4.8%) | 3 (11.5%) |  | 0.151 | -- | 3 (20.0%) | 4.87 (0.81, 20.58) | **0.041** | **0.859** | 0 (0.0%) | -- | -- | -- |
| HLA-C*02:10 | 49 (14.0%) | 5 (19.2%) | 1.47 (36.78, 12.30) | 0.398 | -- | 3 (20.0%) | 1.54 (0.27, 5.99) | 0.467 | -- | 2 (18.2%) | -- | -- | -- |
| HLA-C*04:01 | 81 (23.1%) | 9 (34.6%) | 1.76 (9.13, 11.27) | 0.231 | -- | 4 (26.7%) | 1.21 (0.27, 4.23) | 0.757 | -- | 5 (45.5%) | 2.77 (0.65, 11.20) | 0.141 | -- |
| HLA-C*06:02 | 114 (32.5%) | 9 (34.6%) | 1.10 (37.13, 9.23) | 0.830 | -- | 6 (40.0%) | 1.38 (0.40, 4.48) | 0.579 | -- | 3 (27.3%) | 0.78 (0.13, 3.33) | 1 | -- |
| HLA-C*07:01 | 63 (17.9%) | 5 (19.2%) | 1.09 (39.92, 12.28) | 0.796 | -- | 3 (20.0%) | 1.14 (0.20, 4.41) | 0.740 | -- | 2 (18.2%) | -- | -- | -- |
| HLA-C*17:01 | 51 (14.5%) | 7 (26.9%) | 2.16 (7., 12.30) | 0.096 | -- | 3 (20.0%) | 1.47 (0.26, 5.71) | 0.472 | -- | 4 (36.4%) | 3.35 (0.69, 13.73) | 0.069 | -- |
| HLA-C*18:01 | 14 (4.0%) | 4 (15.4%) | 4.35 (24., 12.33) | **0.028** | 0.791 | 3 (20.0%) | 5.96 (0.97, 25.88) | **0.027** | **0.714** | 1 (9.1%) | -- | -- | -- |

**Table E9.** HLA class I genotypes of co-trimoxazole-induced SCAR patients in Australia. HLA alleles are highlighted according to alignments with shared peptide binding specificities (B*44:03-like, green; B*38:01-like, blue) or carriage of HLA-C*04:01 (orange). *F, female; M, male*.

| Patient | Culprit drug | Sex | HLA-A<br>allele 1 | HLA-A<br>allele 2 | HLA-B<br>allele 1 | HLA-B<br>allele 2 | HLA-C<br>allele 1 | HLA-C<br>allele 2 | B*44:03-like<br>cluster | B*38:01-like<br>cluster | C*04:01<br>allele |
| --- | --- | --- | --- | --- | --- | --- | --- | --- | --- | --- | --- |
| AUSSJS/TEN 1 | Co-trimoxazole | F | A*11:02 | A*33:03 | B*15:02 | B*44:03 | C*07:01 | C*08:01 |  |  |  |
| AUSSJS/TEN 2 | Co-trimoxazole | F | A*24:02 | A*34:01 | B*13:01 | B*56:02 | C*01:02 | C*12:03 |  |  |  |
| AUSSJS/TEN 3 | Co-trimoxazole | M | A*03:01 | A*26:01 | B*07:02 | B*44:02 | C*07:04 | C*12:03 |  |  |  |
| AUSSJS/TEN 4 | Co-trimoxazole | F | A*02:01 | A*02:01 | B*40:01 | B*44:02 | C*03:04 | C*05:01 |  |  |  |
| AUSSJS/TEN 5 | Co-trimoxazole | F | A*02:01 | A*26:01 | B*38:01 | B*44:02 | C*05:01 | C*12:03 |  |  |  |
| AUSSJS/TEN 6 | Co-trimoxazole | M | A*02:01 | A*03:01 | B*07:02 | B*55:01 | C*03:03 | C*07:02 |  |  |  |
| AUSDRESS 1 | Co-trimoxazole | F | A*02:01 | A*02:01 | B*37:01 | B*41:02 | C*06:02 | C*17:01 |  |  |  |
| AUSDRESS 2 | Co-trimoxazole | M | A*25:01 | A*68:01 | B*18:01 | B*51:01 | C*12:03 | C*15:02 |  |  |  |
| AUSDRESS 3 | Co-trimoxazole | M | A*02:01 | A*23:01 | B*15:01 | B*49:01 | C*03:03 | C*07:01 |  |  |  |
| AUSDRESS 4 | Co-trimoxazole | F | A*11:01 | A*11:01 | B*13:01 | B*46:01 | C*01:02 | C*03:04 |  |  |  |
| AUSDRESS 5 | Co-trimoxazole | M | A*01:01 | A*02:11 | B*52:01 | B*57:01 | C*06:02 | C*12:02 |  |  |  |
| AUSDRESS 6 | Co-trimoxazole | M | A*11:01 | A*33:03 | B*15:02 | B*58:01 | C*03:02 | C*08:01 |  |  |  |

